# PH-LLM: Public Health Large Language Models for Infoveillance

**DOI:** 10.1101/2025.02.08.25321587

**Authors:** Xinyu Zhou, Jiaqi Zhou, Chiyu Wang, Qianqian Xie, Kaize Ding, Chengsheng Mao, Yuntian Liu, Zhiyuan Cao, Huangrui Chu, Xi Chen, Hua Xu, Heidi J. Larson, Yuan Luo

## Abstract

**Background:** The effectiveness of public health intervention, such as vaccination and social distancing, relies on public support and adherence. Social media has emerged as a critical platform for understanding and fostering public engagement with health interventions. However, the lack of real-time surveillance on public health issues leveraging social media data, particularly during public health emergencies, leads to delayed responses and suboptimal policy adjustments.

**Methods:** To address this gap, we developed PH-LLM (Public Health Large Language Models for Infoveillance)—a novel suite of large language models (LLMs) specifically designed for real-time public health monitoring. We curated a multilingual training corpus comprising 593,100 instruction-output pairs from 36 datasets, covering 96 public health infoveillance tasks and 6 question-answering datasets based on social media data. PH-LLM was trained using quantized low-rank adapters (QLoRA) and LoRA plus, leveraging Qwen 2.5, which supports 29 languages. The PH-LLM suite includes models of six different sizes: 0.5B, 1.5B, 3B, 7B, 14B, and 32B. To evaluate PH-LLM, we constructed a benchmark comprising 19 English and 20 multilingual public health tasks using 10 social media datasets (totaling 52,158 unseen instruction-output pairs). We compared PH-LLM’s performance against leading open-source models, including Llama-3.1-70B-Instruct, Mistral-Large-Instruct-2407, and Qwen2.5-72B-Instruct, as well as proprietary models such as GPT-4o.

**Findings:** Across 19 English and 20 multilingual evaluation tasks, PH-LLM consistently outperformed baseline models of similar and larger sizes, including instruction-tuned versions of Qwen2.5, Llama3.1/3.2, Mistral, and bloomz, with PH-LLM-32B achieving the state-of-the-art results. Notably, PH-LLM-14B and PH-LLM-32B surpassed Qwen2.5-72B-Instruct, Llama-3.1-70B-Instruct, Mistral-Large-Instruct-2407, and GPT-4o in both English tasks (>=56.0% vs. <= 52.3%) and multilingual tasks (>=59.6% vs. <= 59.1%). The only exception was PH-LLM-7B, with slightly suboptimal average performance (48.7%) in English tasks compared to Qwen2.5-7B-Instruct (50.7%), although it outperformed GPT-4o mini (46.9%), Mistral-Small-Instruct-2409 (45.8%), Llama-3.1-8B-Instruct (45.4%), and bloomz-7b1-mt (27.9%).

**Interpretation:** PH-LLM represents a significant advancement in real-time public health infoveillance, offering state-of-the-art multilingual capabilities and cost-effective solutions for monitoring public sentiment on health issues. By equipping global, national, and local public health agencies with timely insights from social media data, PH-LLM has the potential to enhance rapid response strategies, improve policy-making, and strengthen public health communication during crises and beyond.

**Funding:** This study is supported in part by NIH grants R01LM013337 (YL).

## Introduction

The effectiveness of public health interventions, such as social distancing, COVID-19 testing, and vaccination, hinges on collective support, participation, and adherence, in both physically spaces and virtual platforms. With recent advances in machine learning, infoveillance—the continuous analysis of online text information^1^—has emerged as a supplement to traditional public health surveillance approaches, offering early insights into public responses to interventions. Infoveillance has also been employed to mitigate the infodemic–an overwhelming surge of information and misinformation that may lead to deleterious public health consequences during pandemics, in addition to ensuring public adherence and informing health policy decisions.^1–5^

A growing number of public health researchers and authorities are leveraging social media data to explore vaccine attitudes, mental health issues, adherence to non-pharmaceutical interventions (NPIs), the spread of misinformation, and beyond, with machine learning models such as random forest and naïve bayes.^2,6,7^ Despite these efforts, real-time infoveillance on social media platforms remains limited, especially when tracking rapidly evolving public health emergencies like COVID-19^6,7^. Without timely and scalable infoveillance methods, there may potentially be delays in policy refinement and missed opportunities for prompt public health interventions.^7^

Large Language Models (LLMs) hold promising potentials for infoveillance^8–13^. They can perform infoveillance tasks without necessitating the extensive resources, time, and task-specific annotated datasets typically required for training conventional machine learning models for large-scale infoveillance. Moreover, their human-like interactions make them more accessible to public health experts than many other machine learning tools. However, proprietary LLMs such as ChatGPT are associated with significant cost and may lead to data leakage. On the other hand, open-source LLMs targeted general tasks are not optimized for public health inforveillance. There’s a need for developing LLMs tailored for public health infoveillance, which could significantly reduce costs while delivering state-of-the-art performance.

In this study, we introduce PH-LLM (Public Health Large Language Models for infoveillance), which is a novel suite of LLMs specifically trained for multilingual infoveillance on social media platforms. We designed the first multilingual public health infoveillance benchmark, where we evaluated PH-LLM against leading open-source and proprietary LLMs including GPT-4o. The PH-LLM models and associated Python code can be publicly accessible at https://github.com/luoyuanlab/PH-LLM.

## Methods

In this study, we developed a suite of LLMs named PH-LLM, available in six sizes for various computing settings: PH-LLM-0.5B, PH-LLM-1.5B, PH-LLM-3B, PH-LLM-7B, PH-LLM-14B, and PH-LLM-32B. These PH-LLM models were instruction-based fine-tuned on top of Qwen 2.5,^14^ using a curated dataset of 593,100 instruction-output pairs based on 30 infoveillance datasets with a total of 96 public health infoveillance tasks and six question-answering datasets. We evaluated the PH-LLM models on 39 tasks with a total of 52,158 instruction-output pairs, across 10 datasets in English, Chinese, Arabic, and Indonesian. Notably, the evaluation datasets were distinct from those used during the development of PH-LLM. The performance of PH-LLM was benchmarked against state-of-the-art instruction-tuned LLMs, including GPT-4o (version 2024-05-13), Llama-3.1-70B-Instruct, Mistral-Large-Instruct-2407, and Qwen2.5-72B-Instruct^14–17^. An overview of this study is provided in Figure 1.

**Figure 1.**
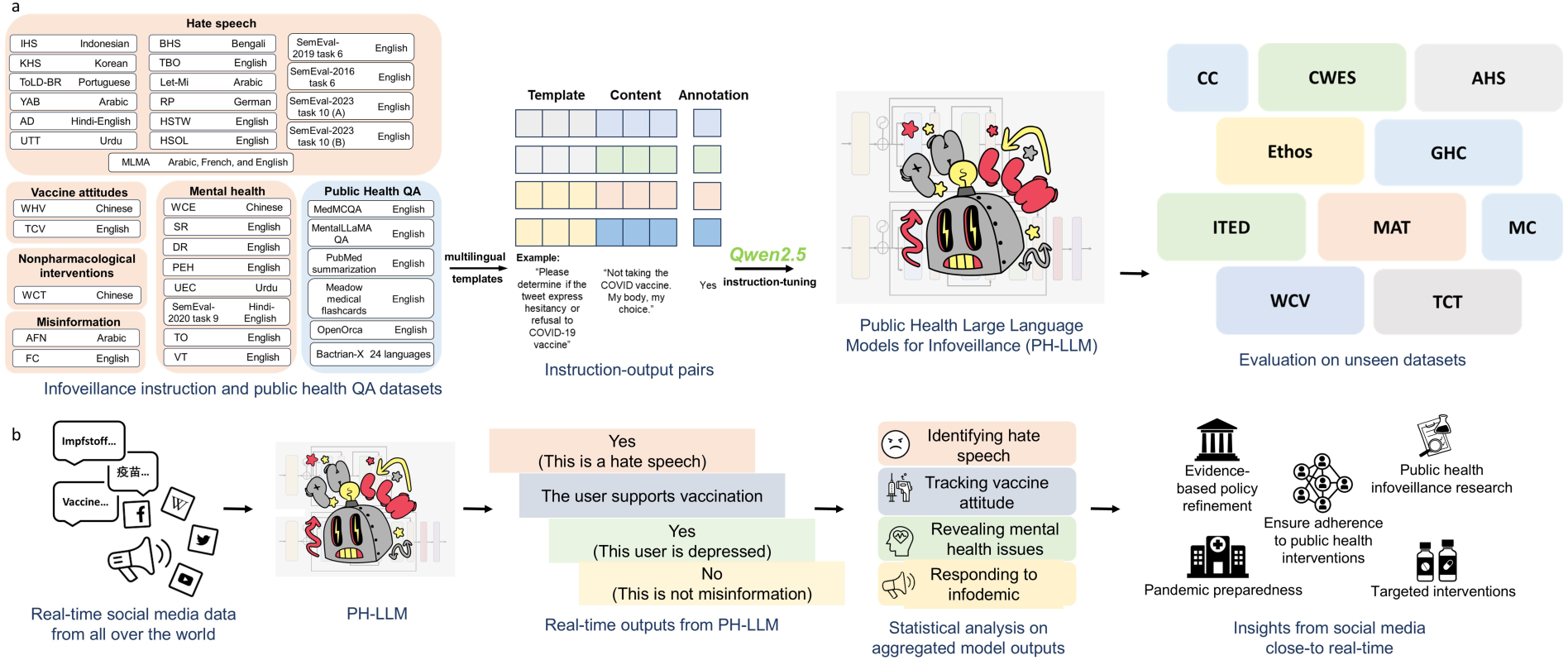
Overview of this study.

### Data Source

We conducted a Google search to compile an initial list of publicly available, manually annotated infoveillance datasets based on social media data. Two researchers (XZ and JZ, or XZ and CW) assessed the annotation quality of each dataset. The evaluation criteria included subjective impressions of the study’s quality, the robustness of the annotation process described, and the popularity of the paper and dataset as indicated by metrics such as citations and GitHub stars. Discrepancies between researchers were resolved through discussions. Only datasets that were manually annotated and deemed high quality by both researchers were recruited, either as the training set or the evaluation dataset. We prioritized datasets requiring API access for inclusion in the evaluation set. Datasets annotated using machine learning models were excluded. Ultimately, a total of 40 infoveillance datasets were collected. Of these, 30 datasets, encompassing 96 public health infoveillance tasks concerning vaccine sentiment, hate speech, mental health, NPIs, misinformation, and beyond, were included in the training set. In addition, six additional QA datasets were incorporated to enrich the training corpus. Details of the training set are provided in Supplementary Table 1. The remaining 10 datasets, comprising 39 unseen infoveillance tasks and 52,158 instruction-output pairs, were excluded from model training and reserved for evaluation. Details of the evaluation datasets are presented in Table 1. Importantly, there was no overlap between the training and evaluation sets.

**Table 1.**
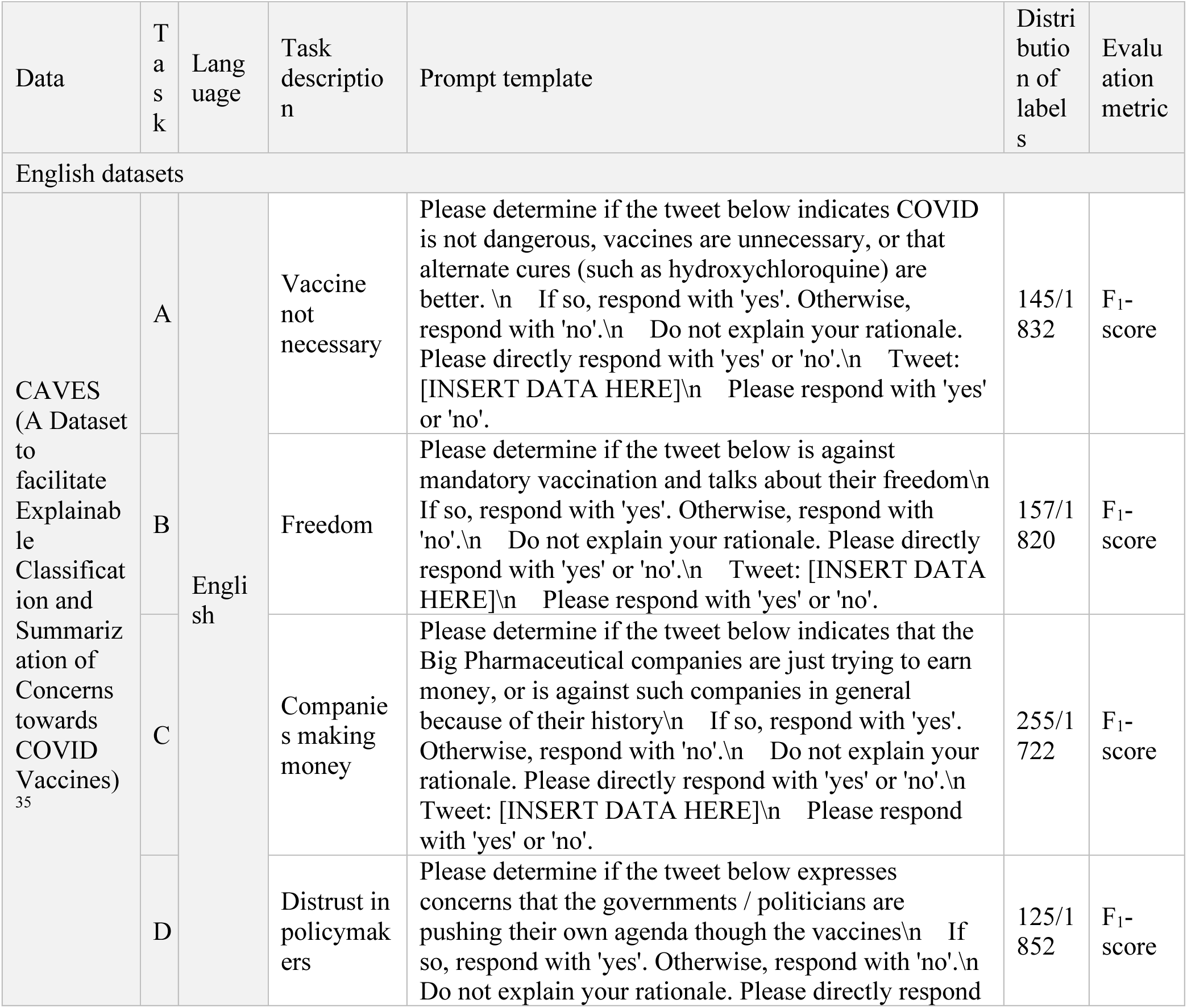
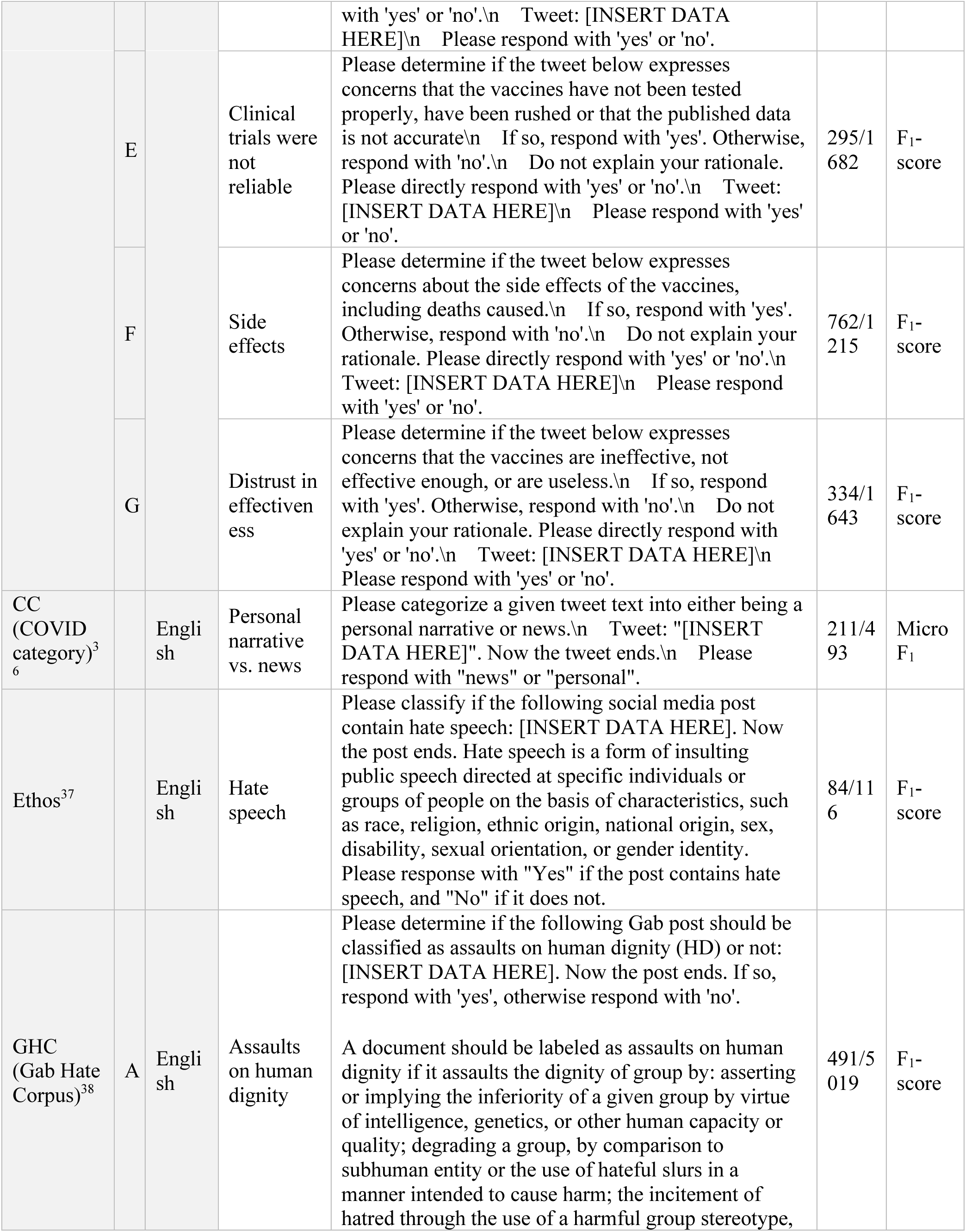
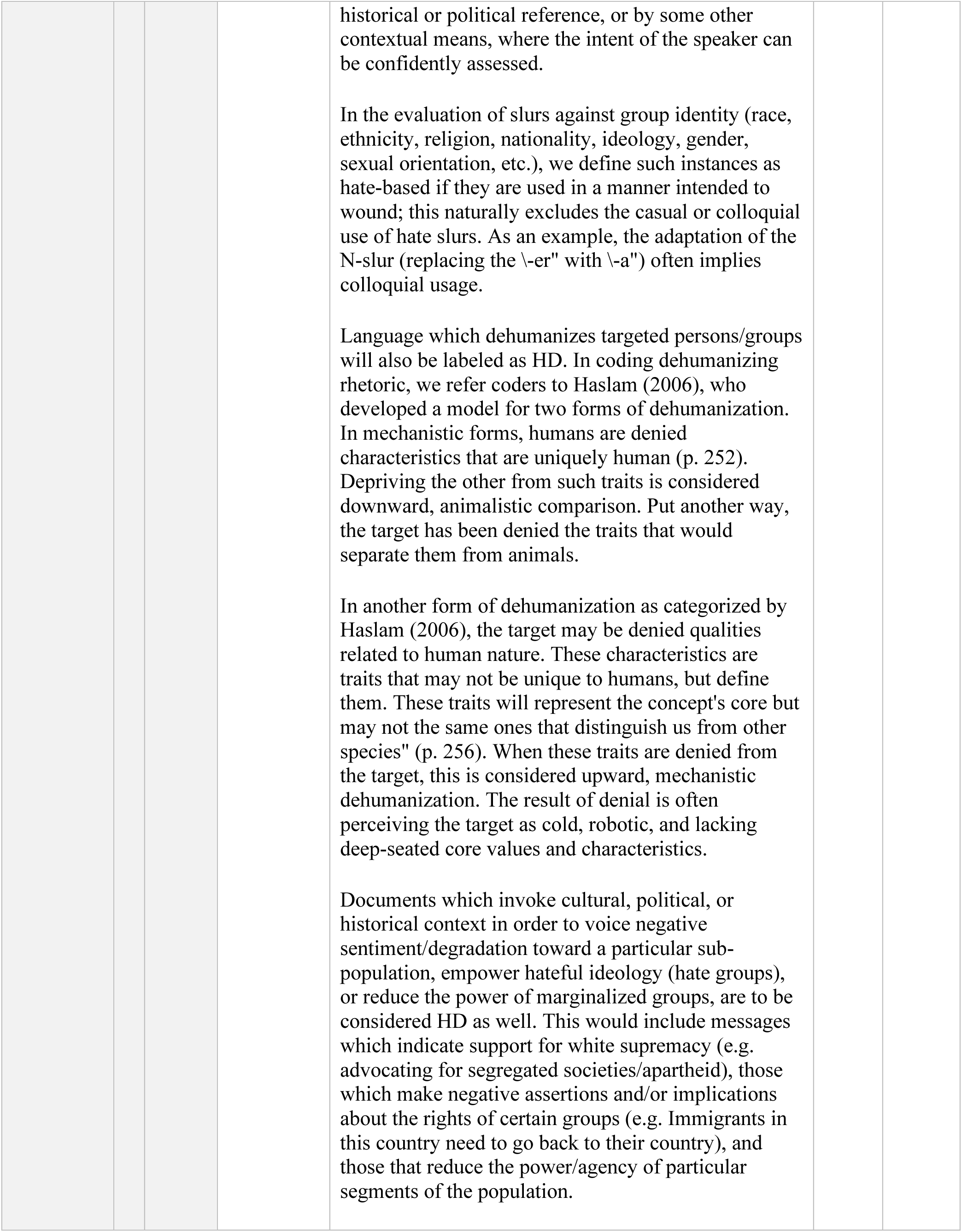
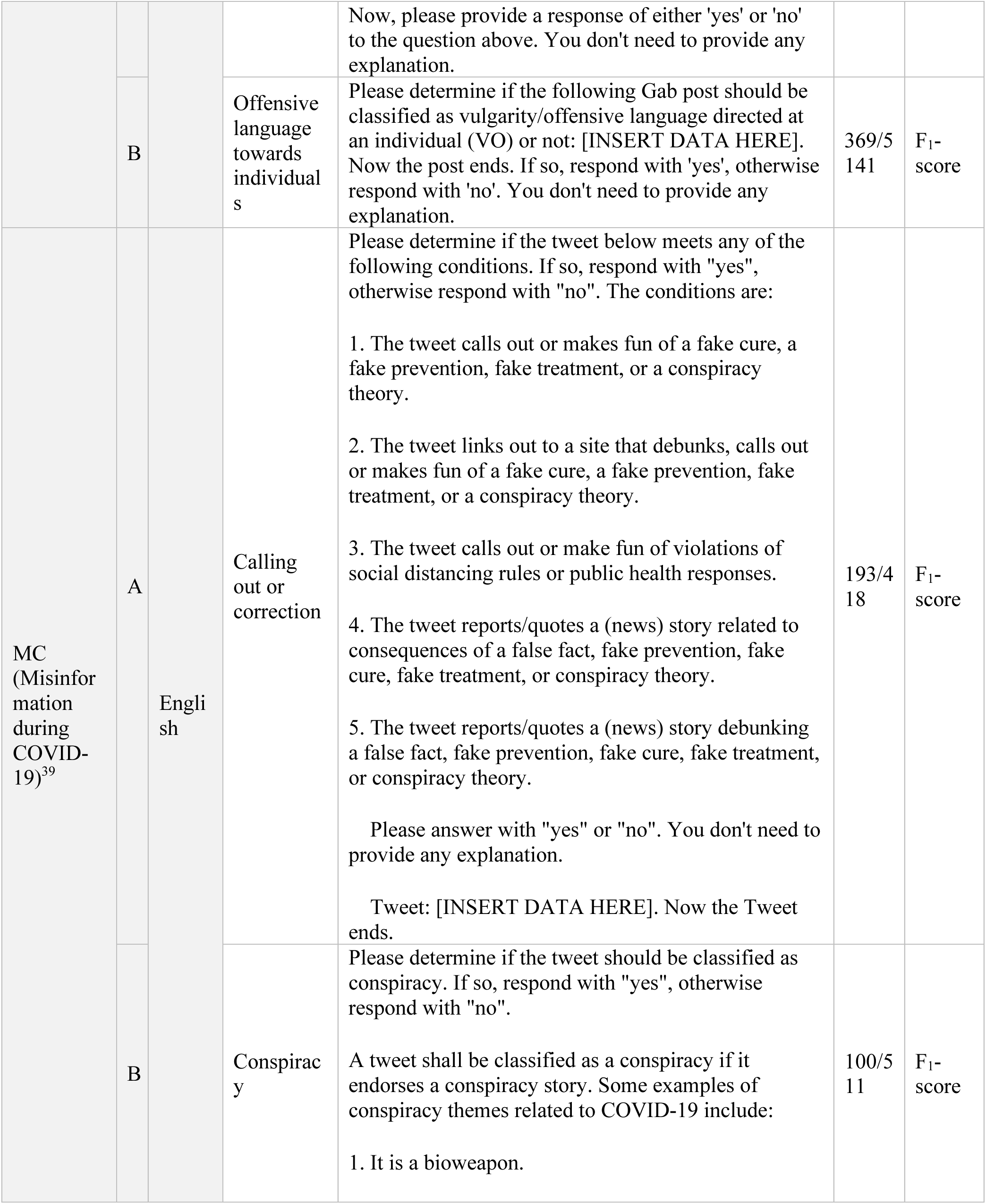
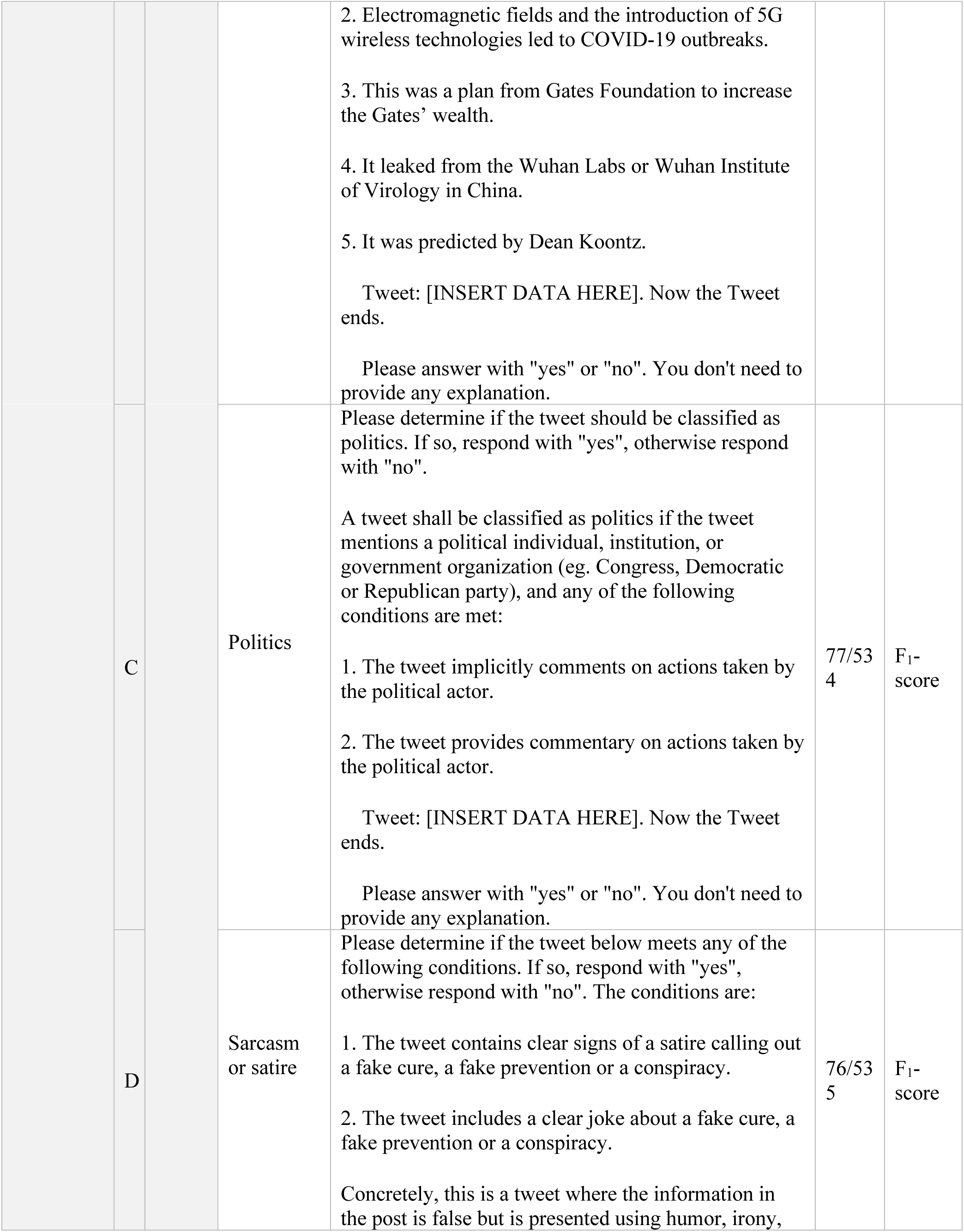
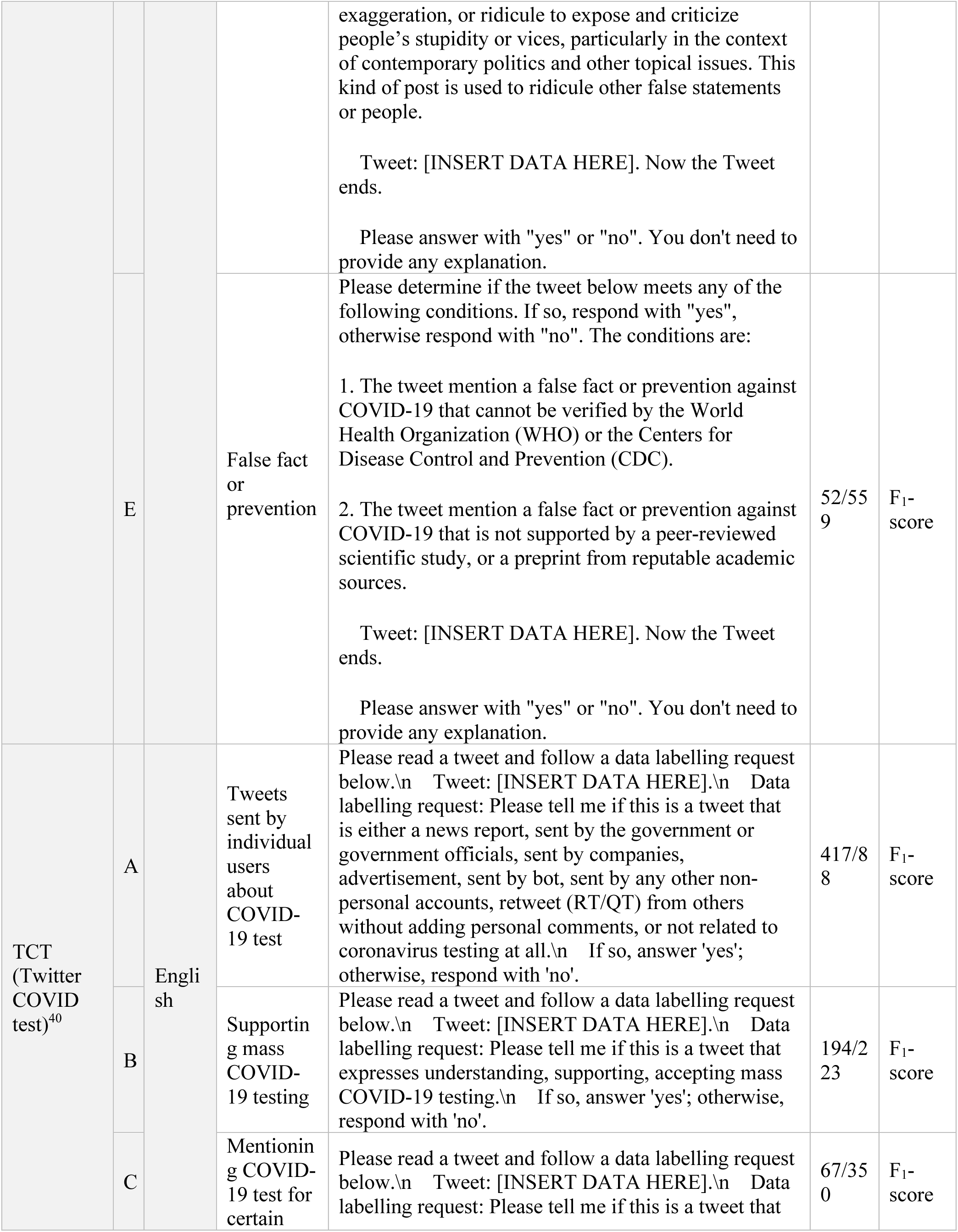
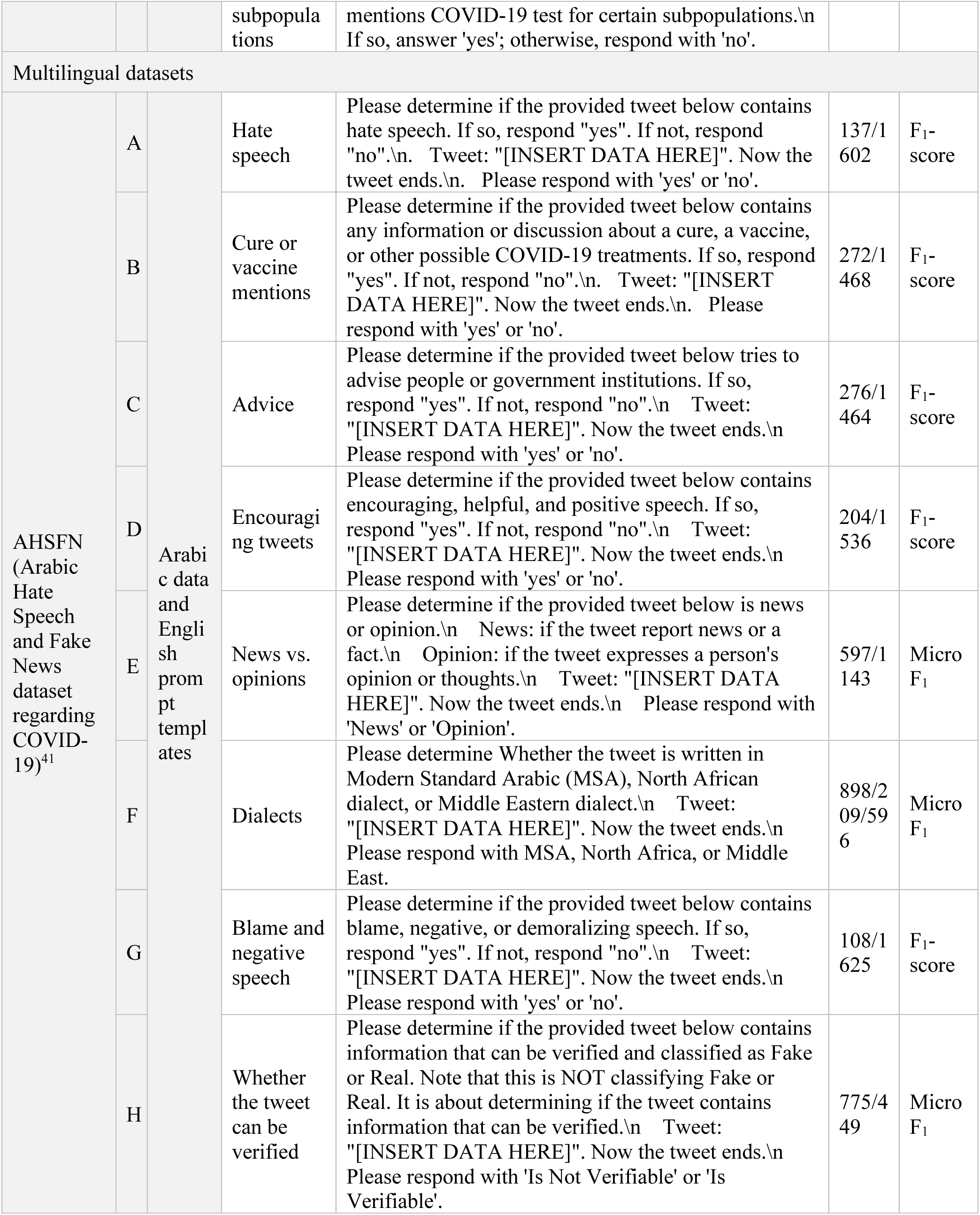
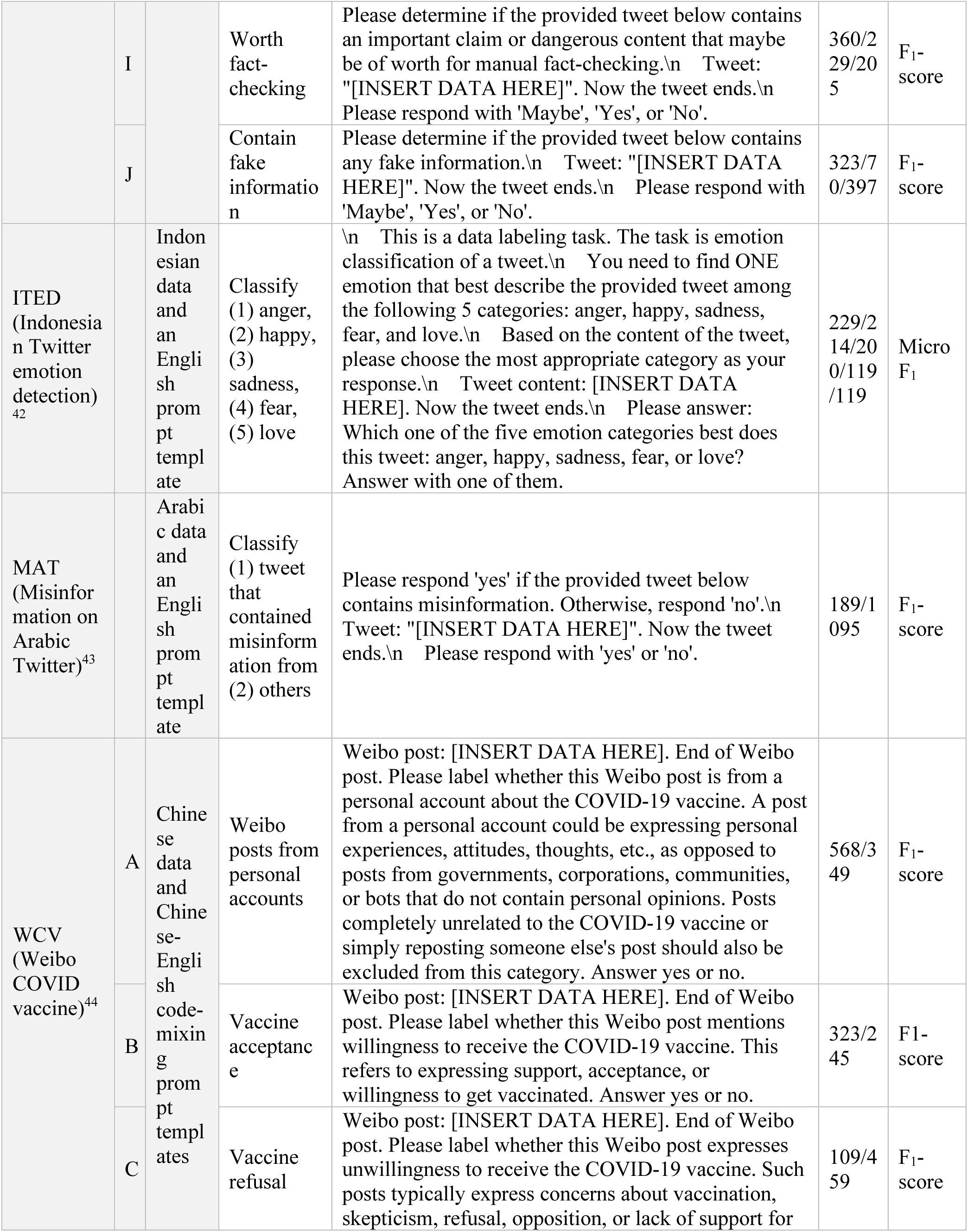
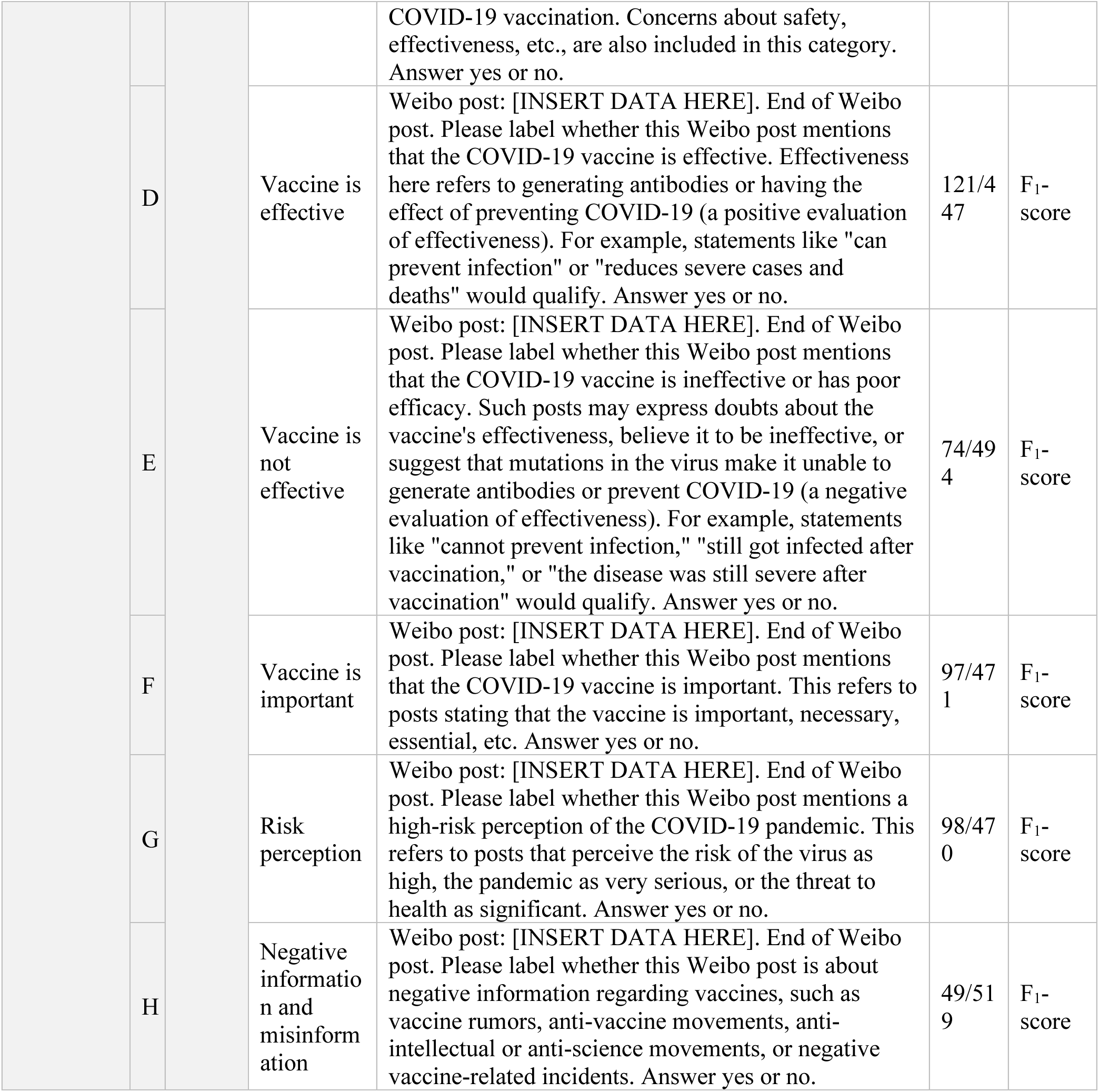
Evaluation benchmark. The evaluation benchmark is based on manually annotated social media datasets. The source of the datasets, language, and the distribution of labels were presented. None of the evaluation datasets were used during the training of PH-LLM models. Note: The prompt templates for the WCV dataset were originally written in Chinese, and their English translations were shown in this table. We construct the prompt templates according to the original data annotation strategy described by the creator of the source datasets without any paraphrasing, whenever possible.

### Constructing Instruction Datasets

#### Infoveillance Instructions (I^2^)

The I^2^ dataset was developed using 30 social media datasets included in the training corpus. Figure 1 illustrated the process of transforming previously annotated social media-based public health infoveillance datasets into instruction datasets, which was applied to create I^2^, an integrated instruction-tuning datasets for training PH-LLM. The datasets in I^2^ were sourced from a total of 30 manually annotated social media datasets from prior studies and were either monolingual or multilingual. Detailed information about each training dataset in I^2^ is provided in Supplementary Table 1. Typically, social media datasets collected from the Internet contain two primary entries: the social media post (or its ID), and corresponding annotation(s) (e.g., 0 and 1). For datasets containing only post IDs (all sourced from X, formally known as Twitter), we retrieved the actual textual content of the post (excluding replies) via the official X API^18^. Each post was transformed into an instruction comprehensible to humans, and its annotation(s) were converted into the gold-standard response for that instruction. Templates were applied to transform social media posts into instructions, as shown in Figure 1.

To enhance the multilingual capabilities of PH-LLM models, templates for developing the I^2^ dataset were translated into 29 languages supported by Qwen 2.5. The list of supported languages is available in Supplementary Material 1.

The original social media posts were not translated. Instead, for each post in I^2^, a template in one of the 29 languages was randomly assigned. Each instruction was a combination of one social media post and one template.^17^ Templates were initially crafted in Chinese or English, and subsequently translated into 28 additional languages using the web interface of ChatGPT-4o (https://chatgpt.com/).

#### Public Health Question Answering (PHQA)

To construct the PHQA dataset, we employed a two-step process. First, we applied a keyword-based filtering approach to extract public health-related instruction-output pairs from three datasets: PubMed summarization, Meadow medical flashcards, and OpenOrca, as a supplement to the training set.^19–21^ The keywords used for filtering are presented in Supplementary Material 1. Second, we selected subsets from the MedMCQA dataset, focusing specifically on two subjects: Social/Preventive Medicine and Psychiatry.^22^ We further sampled 10,000 records from the MentalLLaMA collection, a question-answering dataset regarding mental health derived from gpt-3.5-turbo.^13^ We also supplemented the PHQA with a subset of Bactrian-X,^23^ a multilingual instruction-output dataset generated by gpt-3.5-turbo, to reinforce the multilingual capabilities of our instruction-tuned model.

Merging I^2^ and PHQA yielded our training set consisting of 593,100 instruction-output pairs (Supplement Table 1).

#### Evaluation datasets

For the evaluation benchmark, we selected 10 high-quality, manually annotated social media datasets (Table 1) that were distinct from the training datasets. We included tasks in these datasets where minority classes comprised at least 5% of the data, as extremely imbalance tasks can cause metric fluctuations and may be less relevant to public health. As illustrated in Figure 1, we transformed each record in the evaluation datasets into instruction-output pairs based on prompt templates. The original evaluation datasets, prior to the application of instruction templates, were in English, Chinese, Arabic, or Indonesian, whereas the prompt templates for the evaluation datasets were in English or Chinese-English code-mixing (Table 1). Putting datasets and prompt templates together yielded six evaluation datasets with 19 tasks in English and four multilingual evaluation datasets (two in Arabic-English code-mixing, one in Indonesian-English code-mixing, and one in Chinese-English code-mixing), encompassing 20 tasks. In total, we collected 52,158 instruction-output pairs from 39 tasks across 10 datasets in the evaluation benchmark. Detailed prompt templates for each evaluation task are shown in Table 1.

### Instruction-tuning of the PH-LLM Models

Qwen-2.5, a foundation model developed by Alibaba, was pretrained using up to 18 trillion tokens across more than 29 languages. The model family includes both base model (pretrained only), and instruction models (further trained through instruction-tuning and other methods).^14^ We chose the instruction-tuned version of Qwen2.5 as the backbone for the PH-LLM models, given its multilingual capabilities and superior performance in following human instructions^14^. Supporting over 29 languages, Qwen 2.5 enhances the potential applicability of PH-LLM in global health contexts, including low- and middle-income countries (LMICs).

To enable efficient LLM finetuning, we utilized quantized low-rank adaption (QLoRA).^24,25^ For instruction-tuning, our models were trained over 3 epochs, with an effective batch size of 256, a cut-off length of 1024 tokens, a learning rate of 0.00005, incorporating cosine annealing with a warm-up ratio of 0.1, and LoRAPlus learning rate ratio of 16.^26^ The instruction-tuning process also adhered to Qwen 2.5’s prompt format to maintain consistency. Elaborations on instruction tuning are available in Supplementary Material 1.

### Model Evaluation

We evaluated the zero-shot performance of PH-LLM models—PH-LLM-0.5B, PH-LLM-1.5B, PH-LLM-3B, PH-LLM-7B, PH-LLM-14B, and PH-LLM-32B—against a wide array of open-source and proprietary LLMs, including GPT-4o (version 2024-05-13), Llama-3.1-72B-Instruct, Mistral-Large-Instruct-2407, and Qwen2.5-72B-Instruct.^14–17^ During the evaluation of the open-source models (PH-LLM, Llama, Mistral, BLOOMZ, and Qwen 2.5), we used 4-bit quantization with QLoRA to enhance computational efficiency, using gated GPUs servers at Northwestern University. GPT-4o (version 2024-05-13) and GPT-4o mini (version 2024-07-18) were deployed on the Microsoft Azure platform.

### Statistical Analysis

All evaluation datasets focused on classification task, which represent the predominant type of annotated datasets in public health infoveillance. For classification tasks where only one category was relevant to public health, model performance was assessed using the F_1_ − score. For tasks involving multiple categories of public health significance, we reported the micro F_1_ − score to account for class imbalance. The formulas for calculating precision, recall, F_1_ − score, and micro F_1_ − score are presented in Supplementary Material 1.

## Results

Table 2 compares the zero-shot performance of PH-LLM on 19 tasks across six English-language datasets against other open-source LLMs of similar sizes. PH-LLM models demonstrated superior average performance, as measured by F_1_ − score and micro F_1_ − score, compared to their counterparts. Specifically, the smallest model, PH-LLM-0.5B, achieved an average model performance of 30.3%, outperforming Qwen2.5-0.5B-Instruct (23.6%) across 14 of 19 tasks. PH-LLM-1.5B achieved 39.9%, surpassing both Qwen2.5-1.5B-Instruct (36.3%) and the similar-sized Llama-3.2-1B-Instruct (28.0%) on 15 and 16 out of 19 tasks, respectively.

**Table 2.**
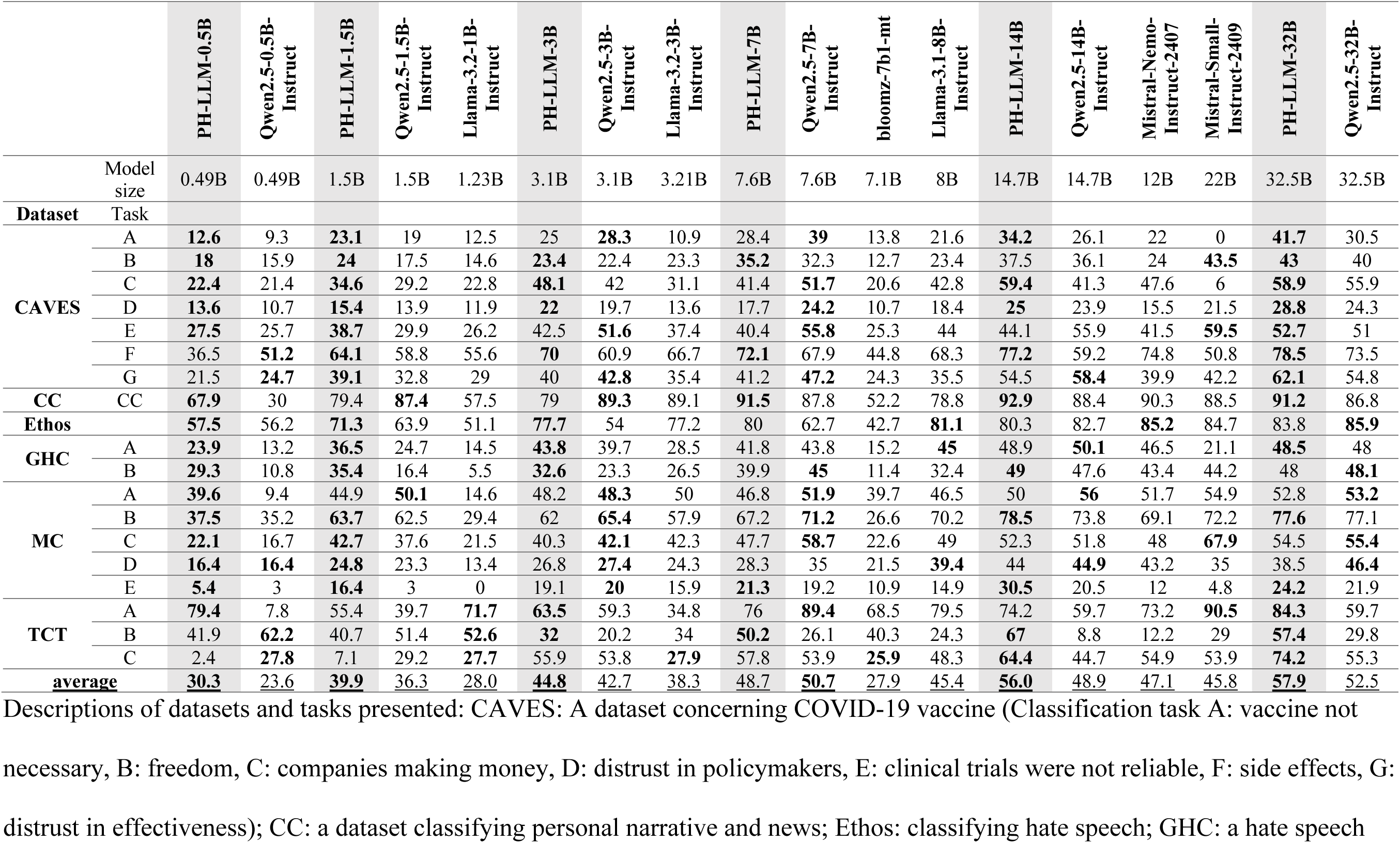
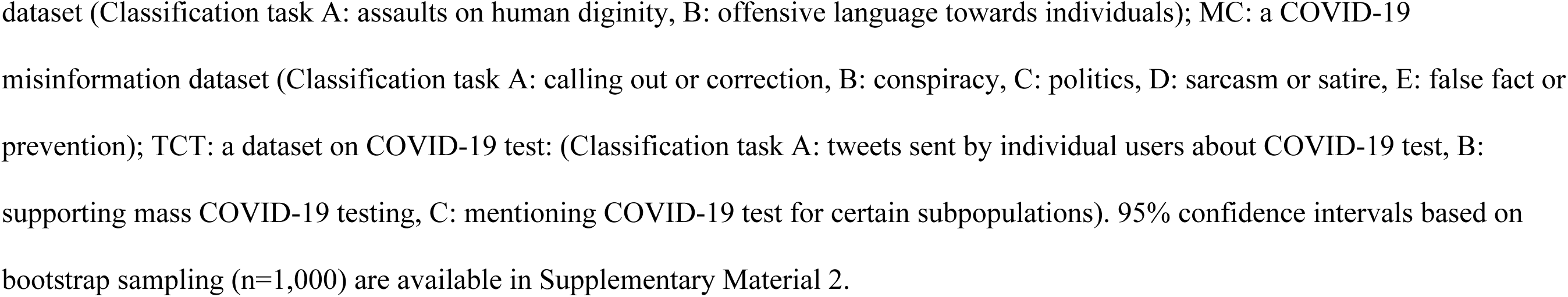
Comparison of zero-shot performance on English-language datasets between PH-LLM and other open-source LLMs of similar sizes.

|  |  | PH-LLM-0.5B | Qwen2.5-0.5B-Instruct | PH-LLM-1.5B | Qwen2.5-1.5B-Instruct | Llama-3.2-1B-Instruct | PH-LLM-3B | Qwen2.5-3B-Instruct | Llama-3.2-3B-Instruct | PH-LLM-7B | Qwen2.5-7B-Instruct | bloomz-7b1-mt | Llama-3.1-8B-Instruct | PH-LLM-14B | Qwen2.5-14B-Instruct | Mistral-Nemo-Instruct-2407 | Mistral-Small-Instruct-2409 | PH-LLM-32B | Qwen2.5-32B-Instruct |
| --- | --- | --- | --- | --- | --- | --- | --- | --- | --- | --- | --- | --- | --- | --- | --- | --- | --- | --- | --- |
|  | Model size | 0.49B | 0.49B | 1.5B | 1.5B | 1.23B | 3.1B | 3.1B | 3.21B | 7.6B | 7.6B | 7.1B | 8B | 14.7B | 14.7B | 12B | 22B | 32.5B | 32.5B |
| **Dataset** | **Task** |  |  |  |  |  |  |  |  |  |  |  |  |  |  |  |  |  |  |
| **CAVES** | A | **12.6** | 9.3 | **23.1** | 19 | 12.5 | 25 | **28.3** | 10.9 | 28.4 | **39** | 13.8 | 21.6 | **34.2** | 26.1 | 22 | 0 | **41.7** | 30.5 |
|  | B | **18** | 15.9 | **24** | 17.5 | 14.6 | **23.4** | 22.4 | 23.3 | **35.2** | 32.3 | 12.7 | 23.4 | 37.5 | 36.1 | 24 | **43.5** | **43** | 40 |
|  | C | **22.4** | 21.4 | **34.6** | 29.2 | 22.8 | **48.1** | 42 | 31.1 | 41.4 | **51.7** | 20.6 | 42.8 | **59.4** | 41.3 | 47.6 | 6 | **58.9** | 55.9 |
|  | D | **13.6** | 10.7 | **15.4** | 13.9 | 11.9 | **22** | 19.7 | 13.6 | 17.7 | **24.2** | 10.7 | 18.4 | **25** | 23.9 | 15.5 | 21.5 | **28.8** | 24.3 |
|  | E | **27.5** | 25.7 | **38.7** | 29.9 | 26.2 | 42.5 | **51.6** | 37.4 | 40.4 | **55.8** | 25.3 | 44 | 44.1 | 55.9 | 41.5 | **59.5** | **52.7** | 51 |
|  | F | 36.5 | **51.2** | **64.1** | 58.8 | 55.6 | **70** | 60.9 | 66.7 | **72.1** | 67.9 | 44.8 | 68.3 | **77.2** | 59.2 | 74.8 | 50.8 | **78.5** | 73.5 |
|  | G | 21.5 | **24.7** | **39.1** | 32.8 | 29 | 40 | **42.8** | 35.4 | 41.2 | **47.2** | 24.3 | 35.5 | 54.5 | **58.4** | 39.9 | 42.2 | **62.1** | 54.8 |
| **CC** | CC | **67.9** | 30 | 79.4 | **87.4** | 57.5 | 79 | **89.3** | 89.1 | **91.5** | 87.8 | 52.2 | 78.8 | **92.9** | 88.4 | 90.3 | 88.5 | **91.2** | 86.8 |
| **Ethos** |  | **57.5** | 56.2 | **71.3** | 63.9 | 51.1 | **77.7** | 54 | 77.2 | 80 | 62.7 | 42.7 | **81.1** | 80.3 | 82.7 | **85.2** | 84.7 | 83.8 | **85.9** |
| **GHC** | A | **23.9** | 13.2 | **36.5** | 24.7 | 14.5 | **43.8** | 39.7 | 28.5 | 41.8 | 43.8 | 15.2 | **45** | 48.9 | **50.1** | 46.5 | 21.1 | **48.5** | 48 |
|  | B | **29.3** | 10.8 | **35.4** | 16.4 | 5.5 | **32.6** | 23.3 | 26.5 | 39.9 | **45** | 11.4 | 32.4 | **49** | 47.6 | 43.4 | 44.2 | 48 | **48.1** |
| **MC** | A | **39.6** | 9.4 | 44.9 | **50.1** | 14.6 | 48.2 | **48.3** | 50 | 46.8 | **51.9** | 39.7 | 46.5 | 50 | **56** | 51.7 | 54.9 | 52.8 | **53.2** |
|  | B | **37.5** | 35.2 | **63.7** | 62.5 | 29.4 | 62 | **65.4** | 57.9 | 67.2 | **71.2** | 26.6 | 70.2 | **78.5** | 73.8 | 69.1 | 72.2 | **77.6** | 77.1 |
|  | C | **22.1** | 16.7 | **42.7** | 37.6 | 21.5 | 40.3 | **42.1** | 42.3 | 47.7 | **58.7** | 22.6 | 49 | 52.3 | 51.8 | 48 | **67.9** | 54.5 | **55.4** |
|  | D | **16.4** | **16.4** | **24.8** | 23.3 | 13.4 | 26.8 | **27.4** | 24.3 | 28.3 | 35 | 21.5 | **39.4** | 44 | **44.9** | 43.2 | 35 | 38.5 | **46.4** |
|  | E | **5.4** | 3 | **16.4** | 3 | 0 | 19.1 | **20** | 15.9 | **21.3** | 19.2 | 10.9 | 14.9 | **30.5** | 20.5 | 12 | 4.8 | **24.2** | 21.9 |
| **TCT** | A | **79.4** | 7.8 | 55.4 | 39.7 | **71.7** | **63.5** | 59.3 | 34.8 | 76 | **89.4** | 68.5 | 79.5 | 74.2 | 59.7 | 73.2 | **90.5** | **84.3** | 59.7 |
|  | B | 41.9 | **62.2** | 40.7 | 51.4 | **52.6** | **32** | 20.2 | 34 | **50.2** | 26.1 | 40.3 | 24.3 | **67** | 8.8 | 12.2 | 29 | **57.4** | 29.8 |
|  | C | 2.4 | **27.8** | 7.1 | 29.2 | **27.7** | 55.9 | 53.8 | **27.9** | 57.8 | 53.9 | **25.9** | 48.3 | **64.4** | 44.7 | 54.9 | 53.9 | **74.2** | 55.3 |
| **average** |  | **30.3** | <u>23.6</u> | **39.9** | <u>36.3</u> | <u>28.0</u> | **44.8** | <u>42.7</u> | <u>38.3</u> | <u>48.7</u> | **50.7** | <u>27.9</u> | <u>45.4</u> | **56.0** | <u>48.9</u> | <u>47.1</u> | <u>45.8</u> | **57.9** | <u>52.5</u> |

Descriptions of datasets and tasks presented: CAVES: A dataset concerning COVID-19 vaccine (Classification task A: vaccine not necessary, B: freedom, C: companies making money, D: distrust in policymakers, E: clinical trials were not reliable, F: side effects, G: distrust in effectiveness); CC: a dataset classifying personal narrative and news; Ethos: classifying hate speech; GHC: a hate speech
dataset (Classification task A: assaults on human dignity, B: offensive language towards individuals); MC: a COVID-19 misinformation dataset (Classification task A: calling out or correction, B: conspiracy, C: politics, D: sarcasm or satire, E: false fact or prevention); TCT: a dataset on COVID-19 test: (Classification task A: tweets sent by individual users about COVID-19 test, B: supporting mass COVID-19 testing, C: mentioning COVID-19 test for certain subpopulations). 95% confidence intervals based on bootstrap sampling (n=1,000) are available in Supplementary Material 2.

Among models with ∼7 billion parameters, PH-LLM-7B achieved 48.7%, outperforming bloomz-7b1-mt (27.9%) and Llama-3.1-8B-Instruct (45.4%). However, it performed slightly below Qwen2.5-7B-Instruct (50.7%). PH-LLM-14B (56.0%) consistently outperformed Qwen2.5-14B-Instruct (48.9%) across 13 out of 19 tasks and exceeded Mistral-Nemo-Instruct-2407 (47.1%) on 17 tasks. Remarkably, it also surpassed Mistral-Small-Instruct-2409 (45.8%), which has a larger parameter size of 22 billion. The largest model, PH-LLM-32B, achieved an average performance of 57.9%, surpassing Qwen2.5-32B-Instruct (52.5%).

Table 3 presents the zero-shot performance of PH-LLM models on 20 tasks across four multilingual datasets with the same set of benchmark LLMs, where PH-LLM consistently outperformed other models of similar sizes. PH-LLM-0.5B improved upon Qwen2.5-0.5B-Instruct (34.5% vs. 29.5%) on 17 out of 20 tasks. Similarly, PH-LLM-1.5B (42.1%) outperformed both Qwen2.5-1.5B-Instruct (34.1%) and Llama-3.2-1B-Instruct (27.7%), while PH-LLM-3B (48.1%) outperformed both Qwen2.5-3B-Instruct (41.1%) and Llama-3.2-3B-Instruct (40.0%). Among models with∼7 billion parameters, PH-LLM-7B (58.5%) consistently outperformed blooms-7b1-mt (27.3%), as well as Qwen2.5-7B-Instruct (47.4%) and Llama-3.1-8B (47.2%) on most of the 20 tasks. PH-LLM-14B (59.6%) also surpassed Qwen2.5-14B-Instruct (51.5%), Mistral-Small-Instruct-2407 (42.9%), and Mistral-Small-Instruct-2409 (47.4%) in most tasks. PH-LLM-32B achieved an average performance of 61.4%, exceeding Qwen2.5-32B-Instruct’s 55.1%.

**Table 3.**
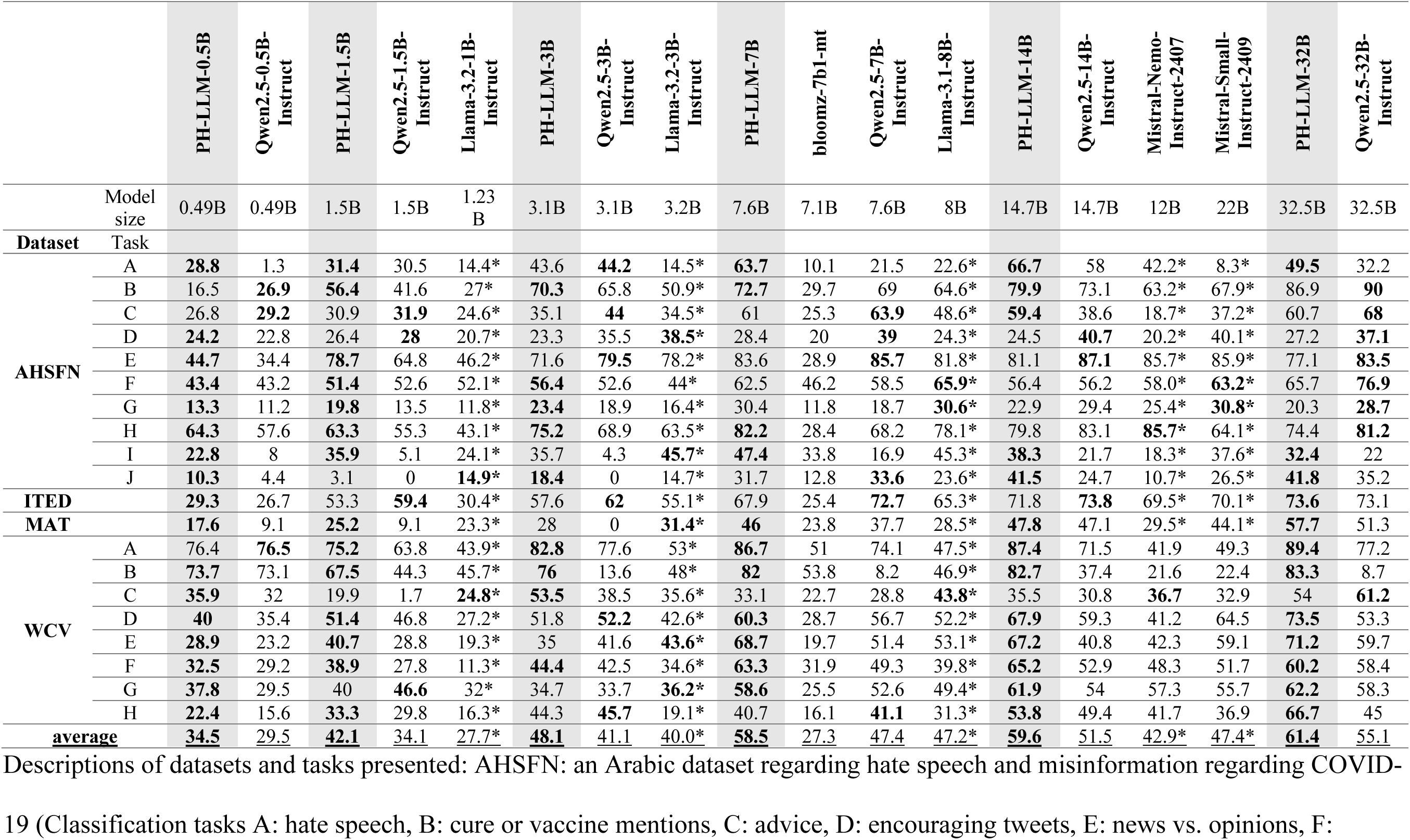
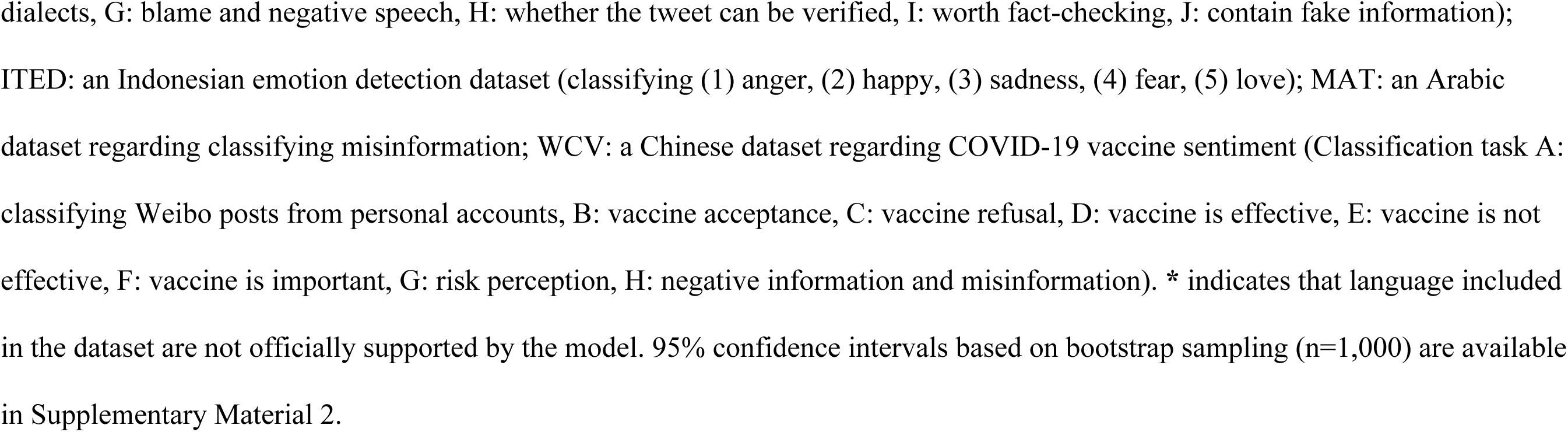
Comparison of zero-shot performance on multilingual datasets between PH-LLM models and other open-source LLMs of similar sizes.

|  |  | PH-LLM-0.5B | Qwen2.5-0.5B-Instruct | PH-LLM-1.5B | Qwen2.5-1.5B-Instruct | Llama-3.2-1B-Instruct | PH-LLM-3B | Qwen2.5-3B-Instruct | Llama-3.2-3B-Instruct | PH-LLM-7B | bloomz-7b1-mt | Qwen2.5-7B-Instruct | Llama-3.1-8B-Instruct | PH-LLM-14B | Qwen2.5-14B-Instruct | Mistral-Nemo-Instruct-2407 | Mistral-Small-Instruct-2409 | PH-LLM-32B | Qwen2.5-32B-Instruct |
| --- | --- | --- | --- | --- | --- | --- | --- | --- | --- | --- | --- | --- | --- | --- | --- | --- | --- | --- | --- |
|  | Model size | 0.49B | 0.49B | 1.5B | 1.5B | 1.23B | 3.1B | 3.1B | 3.2B | 7.6B | 7.1B | 7.6B | 8B | 14.7B | 14.7B | 12B | 22B | 32.5B | 32.5B |
| Dataset | Task |  |  |  |  |  |  |  |  |  |  |  |  |  |  |  |  |  |  |
| AHSFN | A | **28.8** | 1.3 | **31.4** | 30.5 | 14.4* | 43.6 | **44.2** | 14.5* | **63.7** | 10.1 | 21.5 | 22.6* | **66.7** | 58 | 42.2* | 8.3* | **49.5** | 32.2 |
|  | B | 16.5 | **26.9** | **56.4** | 41.6 | 27* | **70.3** | 65.8 | 50.9* | **72.7** | 29.7 | 69 | 64.6* | **79.9** | 73.1 | 63.2* | 67.9* | 86.9 | **90** |
|  | C | 26.8 | **29.2** | 30.9 | **31.9** | 24.6* | 35.1 | **44** | 34.5* | 61 | 25.3 | **63.9** | 48.6* | **59.4** | 38.6 | 18.7* | 37.2* | 60.7 | **68** |
|  | D | **24.2** | 22.8 | 26.4 | **28** | 20.7* | 23.3 | 35.5 | **38.5\*** | 28.4 | 20 | **39** | 24.3* | 24.5 | **40.7** | 20.2* | 40.1* | 27.2 | **37.1** |
|  | E | **44.7** | 34.4 | **78.7** | 64.8 | 46.2* | 71.6 | **79.5** | 78.2* | 83.6 | 28.9 | **85.7** | 81.8* | 81.1 | **87.1** | 85.7* | 85.9* | 77.1 | **83.5** |
|  | F | **43.4** | 43.2 | **51.4** | 52.6 | 52.1* | **56.4** | 52.6 | 44* | 62.5 | 46.2 | 58.5 | **65.9\*** | 56.4 | 56.2 | 58.0* | **63.2\*** | 65.7 | **76.9** |
|  | G | **13.3** | 11.2 | **19.8** | 13.5 | 11.8* | **23.4** | 18.9 | 16.4* | 30.4 | 11.8 | 18.7 | **30.6\*** | 22.9 | 29.4 | 25.4* | **30.8\*** | 20.3 | **28.7** |
|  | H | **64.3** | 57.6 | **63.3** | 55.3 | 43.1* | **75.2** | 68.9 | 63.5* | **82.2** | 28.4 | 68.2 | 78.1* | 79.8 | 83.1 | **85.7\*** | 64.1* | 74.4 | **81.2** |
|  | I | **22.8** | 8 | **35.9** | 5.1 | 24.1* | 35.7 | 4.3 | **45.7\*** | **47.4** | 33.8 | 16.9 | 45.3* | **38.3** | 21.7 | 18.3* | 37.6* | **32.4** | 22 |
|  | J | **10.3** | 4.4 | 3.1 | 0 | **14.9\*** | **18.4** | 0 | 14.7* | 31.7 | 12.8 | **33.6** | 23.6* | **41.5** | 24.7 | 10.7* | 26.5* | **41.8** | 35.2 |
| ITED |  | **29.3** | 26.7 | 53.3 | **59.4** | 30.4* | 57.6 | **62** | 55.1* | 67.9 | 25.4 | **72.7** | 65.3* | 71.8 | **73.8** | 69.5* | 70.1* | **73.6** | 73.1 |
| MAT |  | **17.6** | 9.1 | **25.2** | 9.1 | 23.3* | 28 | 0 | **31.4\*** | **46** | 23.8 | 37.7 | 28.5* | **47.8** | 47.1 | 29.5* | 44.1* | **57.7** | 51.3 |
| WCV | A | 76.4 | **76.5** | **75.2** | 63.8 | 43.9* | **82.8** | 77.6 | 53* | **86.7** | 51 | 74.1 | 47.5* | **87.4** | 71.5 | 41.9 | 49.3 | **89.4** | 77.2 |
|  | B | **73.7** | 73.1 | **67.5** | 44.3 | 45.7* | **76** | 13.6 | 48* | **82** | 53.8 | 8.2 | 46.9* | **82.7** | 37.4 | 21.6 | 22.4 | **83.3** | 8.7 |
|  | C | **35.9** | 32 | 19.9 | 1.7 | **24.8\*** | **53.5** | 38.5 | 35.6* | 33.1 | 22.7 | 28.8 | **43.8\*** | 35.5 | 30.8 | **36.7** | 32.9 | 54 | **61.2** |
|  | D | **40** | 35.4 | **51.4** | 46.8 | 27.2* | 51.8 | **52.2** | 42.6* | **60.3** | 28.7 | 56.7 | 52.2* | **67.9** | 59.3 | 41.2 | 64.5 | **73.5** | 53.3 |
|  | E | **28.9** | 23.2 | **40.7** | 28.8 | 19.3* | 35 | 41.6 | **43.6\*** | **68.7** | 19.7 | 51.4 | 53.1* | **67.2** | 40.8 | 42.3 | 59.1 | **71.2** | 59.7 |
|  | F | **32.5** | 29.2 | **38.9** | 27.8 | 11.3* | **44.4** | 42.5 | 34.6* | **63.3** | 31.9 | 49.3 | 39.8* | **65.2** | 52.9 | 48.3 | 51.7 | **60.2** | 58.4 |
|  | G | **37.8** | 29.5 | 40 | **46.6** | 32* | 34.7 | 33.7 | **36.2\*** | **58.6** | 25.5 | 52.6 | 49.4* | **61.9** | 54 | 57.3 | 55.7 | **62.2** | 58.3 |
|  | H | **22.4** | 15.6 | **33.3** | 29.8 | 16.3* | 44.3 | **45.7** | 19.1* | 40.7 | 16.1 | **41.1** | 31.3* | **53.8** | 49.4 | 41.7 | 36.9 | **66.7** | 45 |
| average |  | **34.5** | **29.5** | **42.1** | **34.1** | **27.7\*** | **48.1** | **41.1** | **40.0\*** | **58.5** | **27.3** | **47.4** | **47.2\*** | **59.6** | **51.5** | **42.9\*** | **47.4\*** | **61.4** | **55.1** |

Descriptions of datasets and tasks presented: AHSFN: an Arabic dataset regarding hate speech and misinformation regarding COVID-19 (Classification tasks A: hate speech, B: cure or vaccine mentions, C: advice, D: encouraging tweets, E: news vs. opinions, F:
dialects, G: blame and negative speech, H: whether the tweet can be verified, I: worth fact-checking, J: contain fake information); ITED: an Indonesian emotion detection dataset (classifying (1) anger, (2) happy, (3) sadness, (4) fear, (5) love); MAT: an Arabic dataset regarding classifying misinformation; WCV: a Chinese dataset regarding COVID-19 vaccine sentiment (Classification task A: classifying Weibo posts from personal accounts, B: vaccine acceptance, C: vaccine refusal, D: vaccine is effective, E: vaccine is not effective, F: vaccine is important, G: risk perception, H: negative information and misinformation). \* indicates that language included in the dataset are not officially supported by the model. 95% confidence intervals based on bootstrap sampling (n=1,000) are available in Supplementary Material 2.

Table 4 and Table 5 presents further comparison of PH-LLM models with larger open-source models and proprietary LLMs for both English-language and multilingual datasets. For English-language comparison (Table 4) across 19 tasks, the largest PH-LLM model, PH-LLM-32B (57.9%), demonstrated not only competitive but superior overall performance to other larger open-source models, such as Qwen2.5-72B-Instruct (49.6%) and Llama-3.1-70B-Instruct (52.3%), and Mistral-Large-Instruct-2407 (51.8%). Furthermore, PH-LLM-32B achieved state-of-the-art performance that it outperformed both proprietary LLMs (46.9% of GPT-4o mini and 50.7% of GPT-4o). In Table 5, for multilingual datasets, PH-LLM-32B continues to outperform all other state-of-the-art models, achieving an average model performance of 61.4% across 20 tasks, specifically Qwen2.5-72B-Instruct (58.5%), Llama-3.1-70B-Instruct (57.7%), Mistral-Large-Instruct-2407 (56.6%), as well as GPT-4o mini (54.1%) and GPT-4o (59.1%).

**Table 4.**
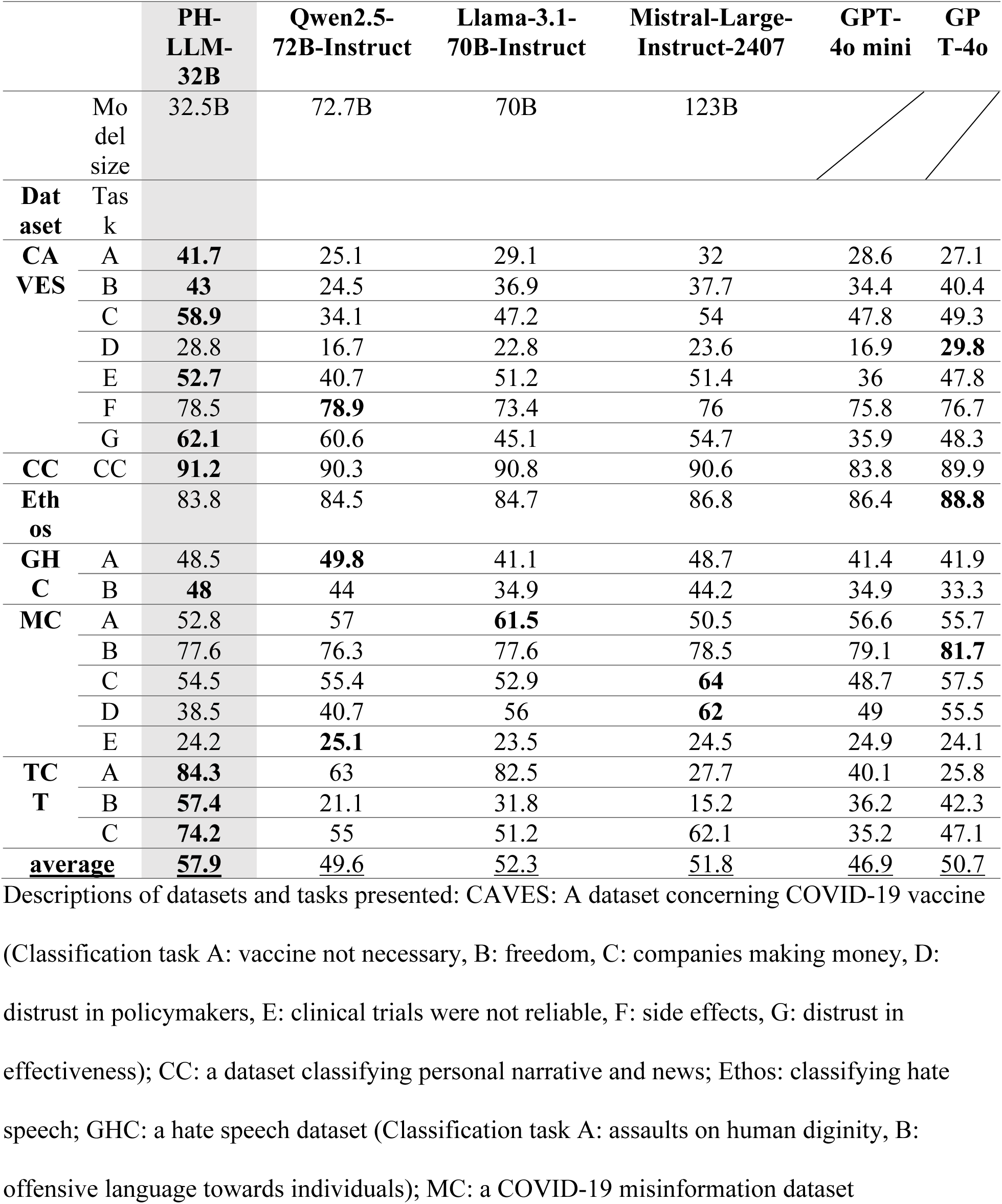
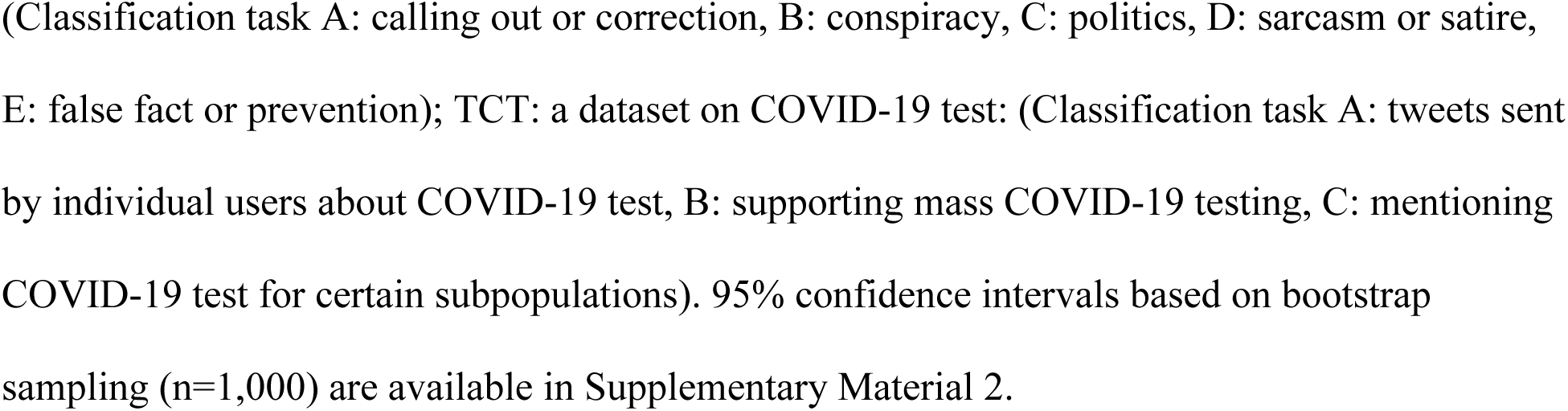
Comparison of zero-shot performance on English-language datasets between PH-LLM-32B and larger open-source models, flagship open-source models, and proprietary LLMs.

|  |  | **PH-LLM-32B** | **Qwen2.5-72B-Instruct** | **Llama-3.1-70B-Instruct** | **Mistral-Large-Instruct-2407** | **GPT-4o mini** | **GP T-4o** |
| --- | --- | --- | --- | --- | --- | --- | --- |
|  | Model size | 32.5B | 72.7B | 70B | 123B |  |  |
| **Dataset** | **Task** |  |  |  |  |  |  |
| **CAVES** | A | **41.7** | 25.1 | 29.1 | 32 | 28.6 | 27.1 |
|  | B | **43** | 24.5 | 36.9 | 37.7 | 34.4 | 40.4 |
|  | C | **58.9** | 34.1 | 47.2 | 54 | 47.8 | 49.3 |
|  | D | 28.8 | 16.7 | 22.8 | 23.6 | 16.9 | **29.8** |
|  | E | **52.7** | 40.7 | 51.2 | 51.4 | 36 | 47.8 |
|  | F | 78.5 | **78.9** | 73.4 | 76 | 75.8 | 76.7 |
|  | G | **62.1** | 60.6 | 45.1 | 54.7 | 35.9 | 48.3 |
| **CC** | CC | **91.2** | 90.3 | 90.8 | 90.6 | 83.8 | 89.9 |
| **Ethos** |  | 83.8 | 84.5 | 84.7 | 86.8 | 86.4 | **88.8** |
| **GHC** | A | 48.5 | **49.8** | 41.1 | 48.7 | 41.4 | 41.9 |
|  | B | **48** | 44 | 34.9 | 44.2 | 34.9 | 33.3 |
| **MC** | A | 52.8 | 57 | **61.5** | 50.5 | 56.6 | 55.7 |
|  | B | 77.6 | 76.3 | 77.6 | 78.5 | 79.1 | **81.7** |
|  | C | 54.5 | 55.4 | 52.9 | **64** | 48.7 | 57.5 |
|  | D | 38.5 | 40.7 | 56 | **62** | 49 | 55.5 |
|  | E | 24.2 | **25.1** | 23.5 | 24.5 | 24.9 | 24.1 |
| **TC T** | A | **84.3** | 63 | 82.5 | 27.7 | 40.1 | 25.8 |
|  | B | **57.4** | 21.1 | 31.8 | 15.2 | 36.2 | 42.3 |
|  | C | **74.2** | 55 | 51.2 | 62.1 | 35.2 | 47.1 |
| **average** |  | **57.9** | 49.6 | 52.3 | 51.8 | 46.9 | 50.7 |

Descriptions of datasets and tasks presented: CAVES: A dataset concerning COVID-19 vaccine (Classification task A: vaccine not necessary, B: freedom, C: companies making money, D: distrust in policymakers, E: clinical trials were not reliable, F: side effects, G: distrust in effectiveness); CC: a dataset classifying personal narrative and news; Ethos: classifying hate speech; GHC: a hate speech dataset (Classification task A: assaults on human dignity, B: offensive language towards individuals); MC: a COVID-19 misinformation dataset
(Classification task A: calling out or correction, B: conspiracy, C: politics, D: sarcasm or satire, E: false fact or prevention); TCT: a dataset on COVID-19 test: (Classification task A: tweets sent by individual users about COVID-19 test, B: supporting mass COVID-19 testing, C: mentioning COVID-19 test for certain subpopulations). 95% confidence intervals based on bootstrap sampling (n=1,000) are available in Supplementary Material 2.

**Table 5.**
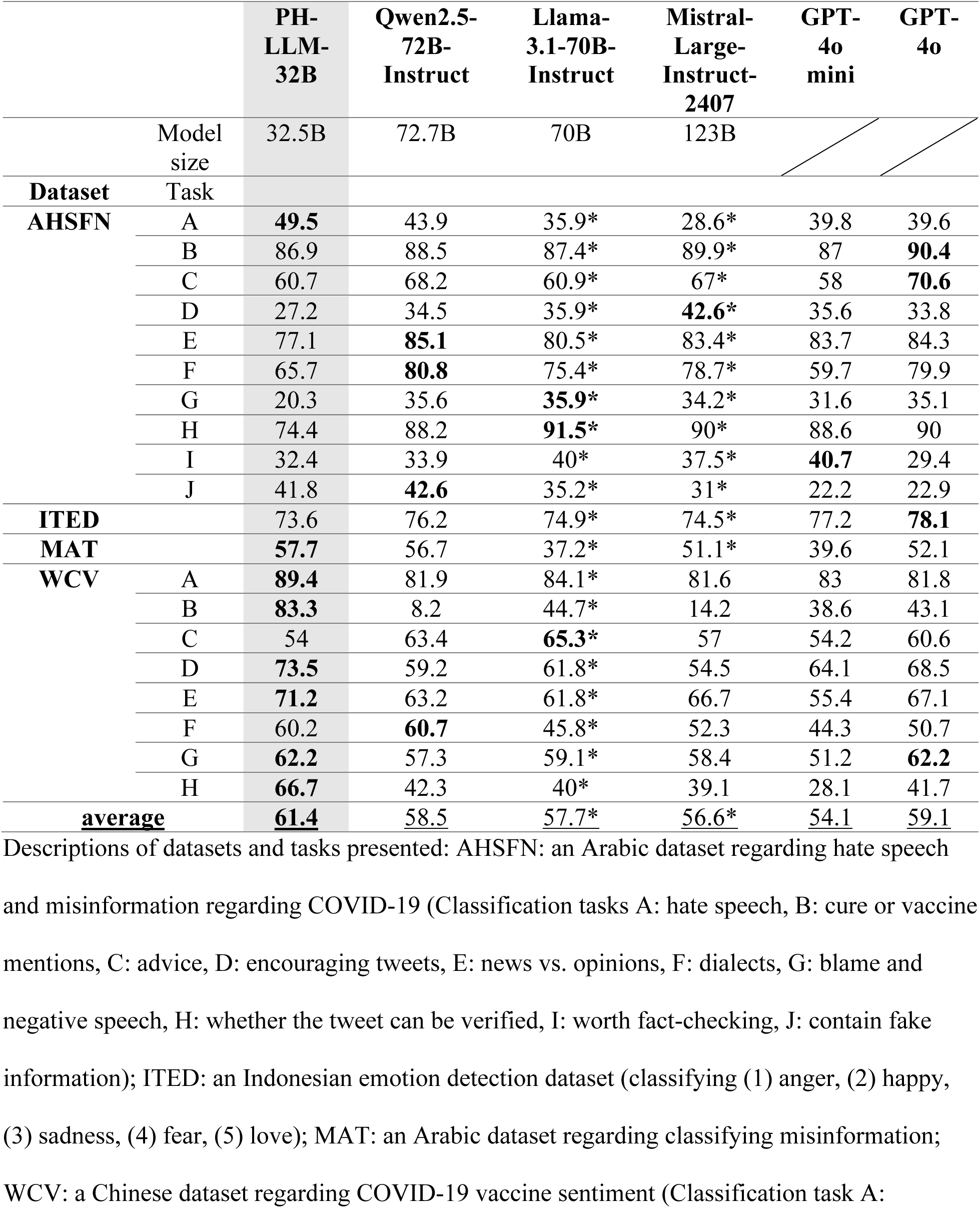
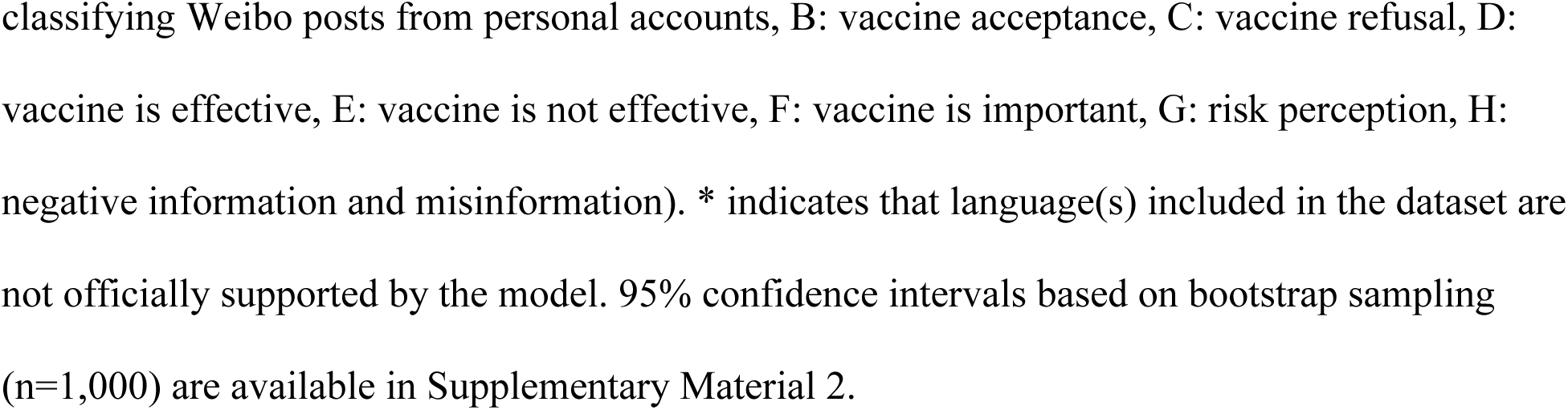
Comparison of zero-shot performance on multilingual datasets between PH-LLM-32B and larger open-source models, flagship open-source models, and proprietary LLMs.

|  |  | **PH-LLM-32B** | **Qwen2.5-72B-Instruct** | **Llama-3.1-70B-Instruct** | **Mistral-Large-Instruct-2407** | **GPT-4o mini** | **GPT-4o** |
| --- | --- | --- | --- | --- | --- | --- | --- |
|  | Model size | 32.5B | 72.7B | 70B | 123B |  |  |
| **Dataset** | **Task** |  |  |  |  |  |  |
| **AHSFN** | A | **49.5** | 43.9 | 35.9* | 28.6* | 39.8 | 39.6 |
|  | B | 86.9 | 88.5 | 87.4* | 89.9* | 87 | **90.4** |
|  | C | 60.7 | 68.2 | 60.9* | 67* | 58 | **70.6** |
|  | D | 27.2 | 34.5 | 35.9* | **42.6\*** | 35.6 | 33.8 |
|  | E | 77.1 | **85.1** | 80.5* | 83.4* | 83.7 | 84.3 |
|  | F | 65.7 | **80.8** | 75.4* | 78.7* | 59.7 | 79.9 |
|  | G | 20.3 | 35.6 | **35.9\*** | 34.2* | 31.6 | 35.1 |
|  | H | 74.4 | 88.2 | **91.5\*** | 90* | 88.6 | 90 |
|  | I | 32.4 | 33.9 | 40* | 37.5* | **40.7** | 29.4 |
|  | J | 41.8 | **42.6** | 35.2* | 31* | 22.2 | 22.9 |
| **ITED** |  | 73.6 | 76.2 | 74.9* | 74.5* | 77.2 | **78.1** |
| **MAT** |  | **57.7** | 56.7 | 37.2* | 51.1* | 39.6 | 52.1 |
| **WCV** | A | **89.4** | 81.9 | 84.1* | 81.6 | 83 | 81.8 |
|  | B | **83.3** | 8.2 | 44.7* | 14.2 | 38.6 | 43.1 |
|  | C | 54 | 63.4 | **65.3\*** | 57 | 54.2 | 60.6 |
|  | D | **73.5** | 59.2 | 61.8* | 54.5 | 64.1 | 68.5 |
|  | E | **71.2** | 63.2 | 61.8* | 66.7 | 55.4 | 67.1 |
|  | F | 60.2 | **60.7** | 45.8* | 52.3 | 44.3 | 50.7 |
|  | G | **62.2** | 57.3 | 59.1* | 58.4 | 51.2 | **62.2** |
|  | H | **66.7** | 42.3 | 40* | 39.1 | 28.1 | 41.7 |
| **average** |  | **61.4** | <u>58.5</u> | <u>57.7*</u> | <u>56.6*</u> | <u>54.1</u> | <u>59.1</u> |

Descriptions of datasets and tasks presented: AHSFN: an Arabic dataset regarding hate speech and misinformation regarding COVID-19 (Classification tasks A: hate speech, B: cure or vaccine mentions, C: advice, D: encouraging tweets, E: news vs. opinions, F: dialects, G: blame and negative speech, H: whether the tweet can be verified, I: worth fact-checking, J: contain fake information); ITED: an Indonesian emotion detection dataset (classifying (1) anger, (2) happy, (3) sadness, (4) fear, (5) love); MAT: an Arabic dataset regarding classifying misinformation; WCV: a Chinese dataset regarding COVID-19 vaccine sentiment (Classification task A:
classifying Weibo posts from personal accounts, B: vaccine acceptance, C: vaccine refusal, D: vaccine is effective, E: vaccine is not effective, F: vaccine is important, G: risk perception, H: negative information and misinformation). \* indicates that language(s) included in the dataset are not officially supported by the model. 95% confidence intervals based on bootstrap sampling (n=1,000) are available in Supplementary Material 2.

Figure 2 shows the relationship between average model performance and model size of LLMs evaluated across 19 English evaluation tasks. A positive relationship was observed between the number of parameters in open-source models and their performance. Notably, PH-LLM models demonstrated superior performance compared to models of similar sizes and even larger counterparts. PH-LLM-14B (56.0%) and PH-LLM-32B (57.9%) outperformed strong baselines, including GPT-4o (50.7%), Mistral-Large-Instruct-2407 (51.8%), and Llama-3.1-70B-Instruct (52.3%).

**Figure 2.**
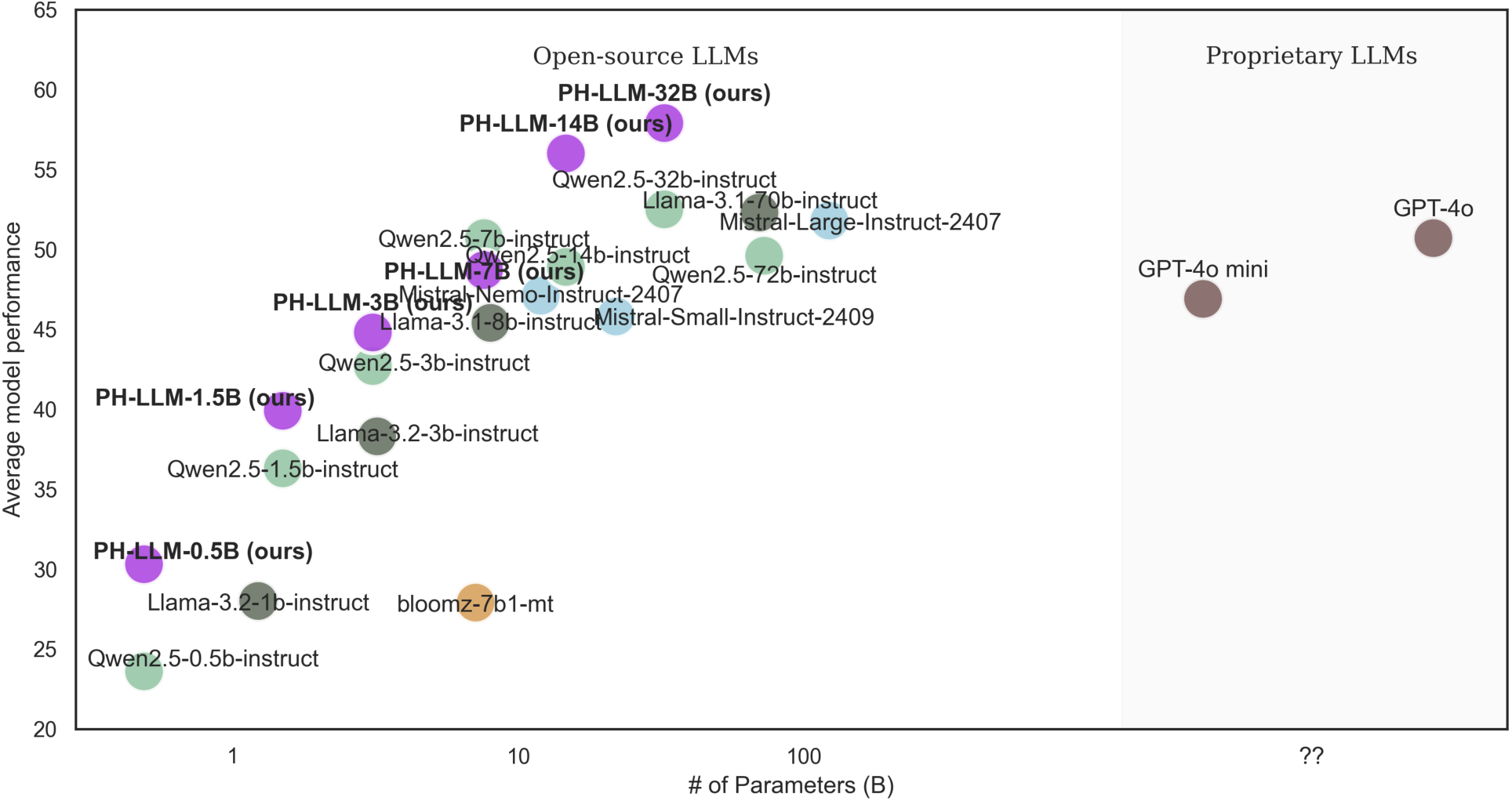
Relationship between model size and average zero-shot performance of LLMs in English evaluation datasets.

Figure 3 shows the relationship between average model performance and model size across 20 multilingual evaluation tasks. PH-LLM consistently outperformed models of similar sizes and in some cases larger models. PH-LLM-7B (58.5%), in particular, matched the average performance as Qwen2.5-72B-Instruct (58.5%). Moreover, both PH-LLM-14B (59.6%) and PH-LLM-32B (61.4%) surpassed state-of-the-art baseline models, including GPT-4o (59.1%), Qwen2.5-72B-Instruct (58.5%), and GPT-4o mini (54.1%).

**Figure 3.**
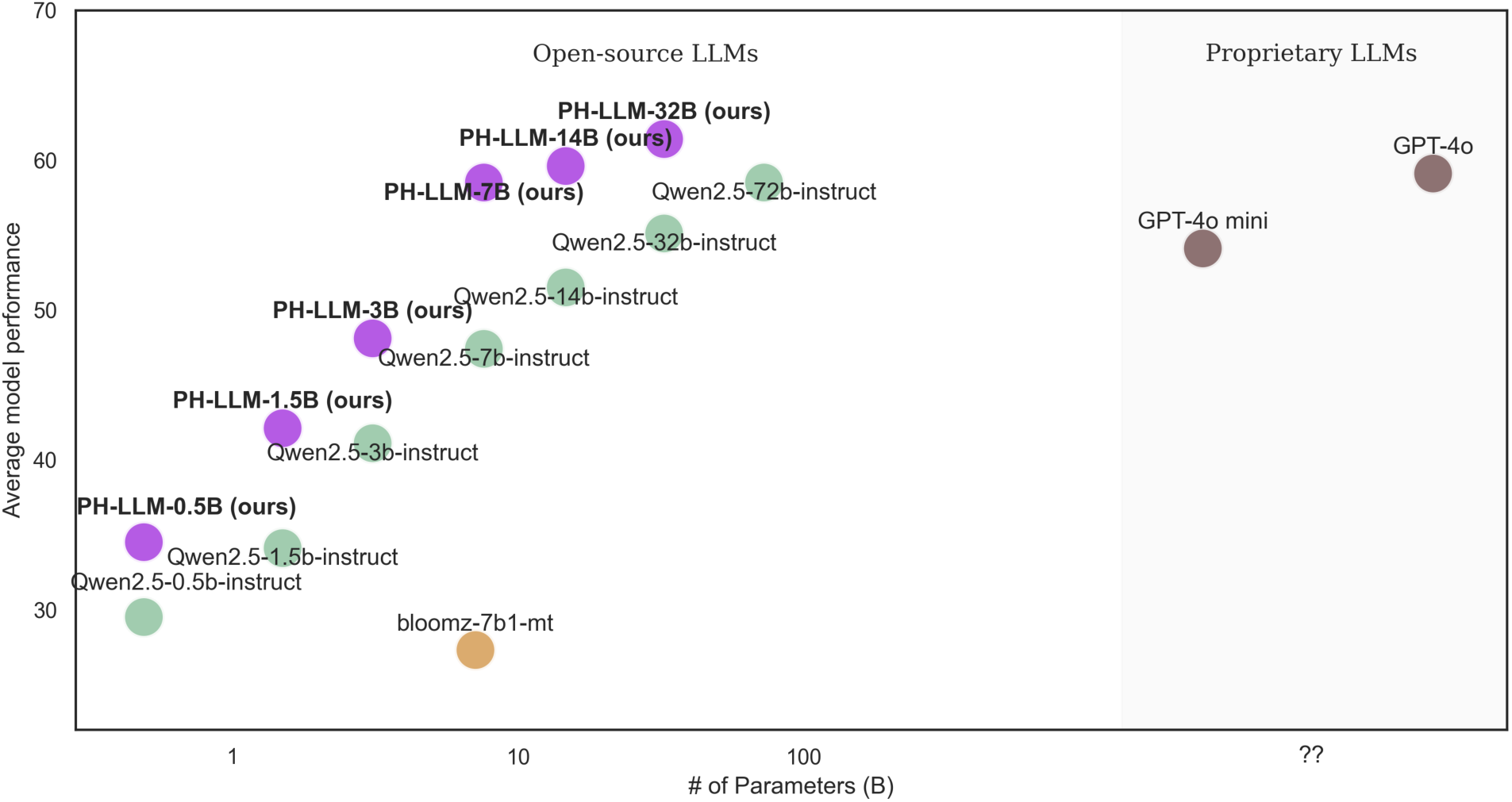
Relationship between model size and average zero-shot performance of LLMs in multilingual evaluation datasets for multilingual LLMs officially supporting all languages in the multilingual evaluation.

## Discussion

In this study, we introduced PH-LLM, a novel suite of LLMs specialized in public health infoveillance. PH-LLM is available in six model sizes: PH-LLM-0.5B, PH-LLM-1.5B, PH-LLM-3B, PH-LLM-7B, PH-LLM-14B, and PH-LLM-32B. Across diverse public health infoveillance tasks, PH-LLM models consistently demonstrated strong performance, outperforming baseline models of comparable or larger sized in most scenarios. Notably, PH-LLM-14B and PH-LLM-32B achieved superior overall performance on 39 tasks from 10 held-out datasets in public health infoveillance settings, surpassing all baseline models including Llama-3.1-72b-instruct, Mistral-Large-Instruct-2407, Qwen2.5-72b-instruct, and GPT-4o.

PH-LLM can reach higher zero-shot performance in public health infoveillance tasks with smaller number of parameters. It reduces the need for extensive GPU resources and complex infrastructure during model deployment and inference, lowering operational costs and making public health infoveillance more accessible, particularly for resource-constrained settings. PH-LLM’s adaptability enables localized and contextualized responses to diverse public health challenges, offering transformative potential for LMICs and other underserved regions.

To the best of our knowledge, PH-LLM is the first suite of LLMs specialized in public health infoveillance which is multilingual and publicly available. Previous studies have utilized general-purpose LLMs to advance public health infoveillance on social media platforms, including tasks like data augmentation in social media datasets,^9,27^ and analyzing public health topics such as vaccine sentiment, mask-wearing behaviors, and mental health.^8,10–13,28^ LLMs have also shown potential in assisting public health practice beyond infoveillance, including pandemic forecasting and information extraction.^29,30^ However, almost all these studies applied general-purpose LLMs like LLaMA and ChatGPT rather than developing LLMs tailored for public health settings,^31,32^ and they focused predominantly on English-language scenarios. PH-LLM emphasizes multilingual capabilities, extending its utility to non-English contexts, which addresses the diverse linguistic needs of global public health.

PH-LLM is designed to be accessible to public health professionals without requiring a background in computer science. With metadata (time, location, social-economic status, and beyond) associated with each social media post, aggregating predictions from PH-LLM can reveal spatiotemporal trends of opinions and behaviors, from nuances on social media platforms, and subsequently underline their public health significance. For example, to inform an HPV vaccination program, public health agencies can apply PH-LLM to stay updated with sudden changes in vaccine acceptance and confidence, trending concerns and misinformation on vaccines, and potential distrust in public health professionals, pharmaceutical companies, or the government. Additionally, tools like LlamaFactory enable users to interact with PH-LLM and effortlessly analyze large-scale data through a user-friendly interface^33^. (Supplementary Figure 1) PH-LLM exhibited strong zero-shot performance for analyzing social media posts relevant to public health. Its performance could be further enhanced potentially through prompt engineering and integration with retrieval-augmented generation and knowledge graph – incorporating contextualized and localized knowledge from public health experts.

PH-LLM equips public health systems with a tool to address future emerging infectious diseases and global health challenges. PH-LLM was trained and evaluated using datasets surrounding vaccine hesitancy, mental health, nonadherence to NPIs, hate speech, and misinformation, and similar challenges may re-emerge in future outbreaks and pandemics.^34^ The generalizability of LLMs also allows PH-LLM to address new and evolving infoveillance topics with greater flexibility towards variations in geographies, languages, populations, and cultural, social, economic and political contexts, which is an advantage over the pretrain-finetune paradigm.

This study has several limitations. First, every LLMs, including PH-LLM, demonstrated suboptimal results in specific tasks. This is because most of the evaluation tasks are imbalanced and could be challenging. Also, we did not optimize prompt templates to ensure fair comparisons and avoid overfitting. Task-specific prompt engineering and evaluations are recommended before deployment of LLMs in zero-shot public health infoveillance. Second, the training set included only 96 infoveillance tasks, which may limit performance of PH-LLM on tasks less represented within the training corpus. Third, PH-LLM’s training datasets were derived from various previous studies, which may reflect inconsistency in annotation quality and potential biases introduced by annotators. Forth, social media data, which underpins PH-LLM’s training and evaluation, represents a biased subset of the population. Predictions based on such data should be interpreted with caution, especially in contexts involving censorship or self-censorship. Lastly, the evaluation focused exclusively on zero-shot performance, and the few-shot and fine-tuning capabilities of PH-LLM remains untested.

Despite these limitations, PH-LLM represents a significant enhancement as a novel suite of LLM tailored for public health infoveillance. Its public availability and state-of-the-art performance demonstrate its potential in public health monitoring and evidence-based policymaking, including in LMICs and among at-risk populations. PH-LLM aspires to equip public health agencies at all levels—global, national and local—with the power of AI to promote public health awareness, inform policy and interventions, and address future global health challenges.

## Data Availability

Models and Python code are available on GitHub (https://github.com/luoyuanlab/PH-LLM). Unfortunately, due to the policy of social media platforms, we cannot share data directly.

## CRediT author statement

Conceptualization: Xinyu Zhou, Yuan Luo; Methodology: Xinyu Zhou, Yuan Luo, Jiaqi Zhou, Chiyu Wang, Kaize Ding, Qianqian Xie, Yuntian Liu, Zhiyuan Cao, Hua Xu; Software: Xinyu Zhou, Chiyu Wang, Jiaqi Zhou, Huangrui Chu; Validation: Jiaqi Zhou; Formal analysis: Xinyu Zhou; Investigation: Xinyu Zhou; Resources: Yuan Luo, Heidi J. Larson, Xinyu Zhou, Huangrui Chu; Data Curation: Xinyu Zhou, Heidi J. Larson; Writing - Original Draft: Xinyu Zhou; Writing - Review & Editing: Xinyu Zhou, Jiaqi Zhou, Chiyu Wang, Qianqian Xie, Kaize Ding, Chengsheng Mao, Yuntian Liu, Zhiyuan Cao, Huangrui Chu, Xi Chen, Hua Xu, Heidi J. Larson, Yuan Luo; Visualization: Xinyu Zhou, Jiaqi Zhou; Supervision: Yuan Luo; Project administration: Yuan Luo, Xinyu Zhou; Funding acquisition: Yuan Luo; All authors have read and approved the manuscript.

## Acknowledgments

This study is supported in part by NIH grants R01LM013337 (YL).

