## supplementary material 1 for "PH-LLM: Public Health Large Language Models for Infoveillance"

**Supplementary Table 1. Instruction tuning datasets for model training**

| Data | Language | Source | Task | Topic | Size |
| --- | --- | --- | --- | --- | --- |
| I^2^ | | | | | |
| Vaccine attitudes | | | | | |
| WHV (Weibo HPV vaccine)^1^ | Chinese | Weibo | Hierarchical classification | HPV vaccine | 23,000 |
| TCV (Twitter COVID-19 vaccine)^2^ | English | Twitter (X) | Hierarchical classification | COVID-19 vaccine | 53,000 |
| Mental health | | | | | |
| WCE (Weibo COVID emotion)^3^ | Chinese | Weibo | 7-class classification | Sentiment analysis | 10,500 |
| SR (Stress – Reddit)^4^ | English | Reddit | Binary classification | Stress detection | 3,000 |
| DR (Depression – Reddit)^5^ | English | Reddit | 4-class classification | Depression detection | 500 |
| PEH (perceived emotions in hurricane)^6^ | English | Twitter (X) | Binary classification | Identify perceived emotions in hurricane | 10,000 |
| UEC (emotion classification in Urdu)^7^ | Urdu | Twitter (X) | Multilabel classification | Emotion classification in Urdu | 6,000 |
| SemEval-2020 task 9^8^ | Hindi-English | Twitter (X) | Binary classification | Sentiment analysis | 12,000 |
| TO (Twitter optimists)^9^ | English | Twitter (X) | 3-class classification | Classify “optimistic,” “pessimistic,” or “neutral” tweets | 6,000 |
| VT (vulgarity on Twitter)^10^ | English | Twitter (X) | 5-class classification | Sentiment analysis | 2,500 |
| Nonpharmacological interventions | | | | | |
| WCT (Weibo COVID test)^11^ | Chinese | Weibo | Hierarchical classification | Public response to COVID-19 test in China | 115,000 |
| Hate speech | | | | | |
| IHS (Indonesian hate speech)^12^ | Indonesian | Twitter (X) | Binary classification | Hate speech and abusive language detection | 10,000 |
| BHS (Bengali hate speech)^13^ | Bengali | YouTube and Facebook | Binary classification | Detect hate speech on YouTube and Facebook about various topics | 20,000 |
| KHS (Korean hate speech)^14^ | Korean | Naver (an entertainment news platform) | Binary classification and 3-class classification | Detect hate speech and gender bias in comments | 8,500 |
| ToLD-BR (toxic language dataset for Brazilian Portuguese)^15^ | Portuguese | Twitter (X) | Binary classification | Detect homophobia, obscene, insult, racism, misogyny, and xenophobia | 11,000 |
| YAB (YouTube anti-social behavior)^16^ | Arabic | YouTube | Binary classification | Detection of offensive language in Arabic YouTube comments | 5,000 |
| AD (aggression detection)^17^ | Hindi-English | Facebook, Twitter (X) | 3-class classification | Aggression detection in Hindi-English Code-Mixed social media | 8,000 |
| UTT (Urdu Threating Tweets)^7^ | Urdu | Twitter (X) | Binary classification | Threatening Tweet in Urdu | 4,000 |
| HSTW (hate speech from Twitter and Whisper) ^18^ | English | Twitter (X), Whisper |  | Hate speech from Whisper and Twitter (X) | 1,000 |
| HSOL (hate speech and offensive language)^19^ | English | Twitter (X) | 3-class classification | Hate Speech and Offensive Language | 48,000 |
| SemEval-2019 task 6 ^20^ | English | Twitter (X) | Hierarchical classification | Offensive language classification | 7,900 |
| TBO (Target-Based Offensive Language Identification)^21^ | English | Twitter (X) | Hierarchical classification | OffensEval 2020 (multilingual offensive language detection) | 3,000 |
| SemEval-2023 task 10 (Subtask B)^22^ | English | Gab and Reddit | 4-class classification | Classification of offensive and aggressive sexism posts | 2,000 |
| Let-Mi (misogynistic language on Arabic Levantine Twitter)^23^ | Arabic | Twitter (X) | Hierarchical classification | Classify misogynistic replies towards popular female journalists’ tweets | 5,500 |
| RP (Rheinische Post)^24^ | German | Rheinische Post (Newspaper) | Binary classification | Classify aggressive comments of news articles | 3,000 |
| SemEval-2016 task 6^25^ | English | Twitter (X) |  | Abortion opinion analysis | 600 |
| SemEval-2023 task 10 (Subtask A)^22^ | English | Gab and Reddit | Binary classification | Binary sexism detection | 10,000 |
| MLMA (multilingual and multi-aspect hate speech analysis)^26^ | Arabic, French, and English | Twitter (X) | Multilabel and multiclass | Classify the hostility type and the target of the tweet | 4,000 |
| Misinformation | | | | | |
| AFN (Arabic fake news)^27^ | Arabic | Twitter (X) | Binary classification | Identify fake news | 1,500 |
| FC (fact-checking for public health claims)^28^ | English | Snopes, Politifact, TruthorFiction, FactCheck, FullFact,  Associated Press, Reuters News, and Health News Review | QA | Public health claims’ fact-checking | 8,000 |
| Public Health QA | | | | | |
| MedMCQA^29^ | English | multiple-choice question answering (MCQA) dataset about real world medical entrance exam questions | QA | Filtered by subject name: “Social & Preventive Medicine” or “Psychiatry” | 15,000 |
| MentalLLaMA QA^30^ | English | gpt-3.5 | QA | Built with gpt-3.5, based on annotated mental health datasets from social media | 10,000 |
| PubMed summarization^31^ | English | PubMed | QA | Generate title based on the abstract of a research paper. Filtered using public health keywords* | 5,000 |
| Meadow medical flashcards^32^ | English | Anki medical curriculum flashcards | QA | Flashcards by medical students to assist learning. Filtered using public health keywords* | 1,400 |
| OpenOrca^33^ | English | gpt-3.5, gpt-4 | QA | A general instruction-tuning dataset. Filtered using public health keywords* | 140,000 |
| Bactrian-X^34^ | 24 languages | gpt-3.5 | QA | Generated using gpt-3.5, using prompts translated from Alpaca and Dolly | 19,200 |

**Supplementary Figure 1. Inference using LlamaFactory^36^**

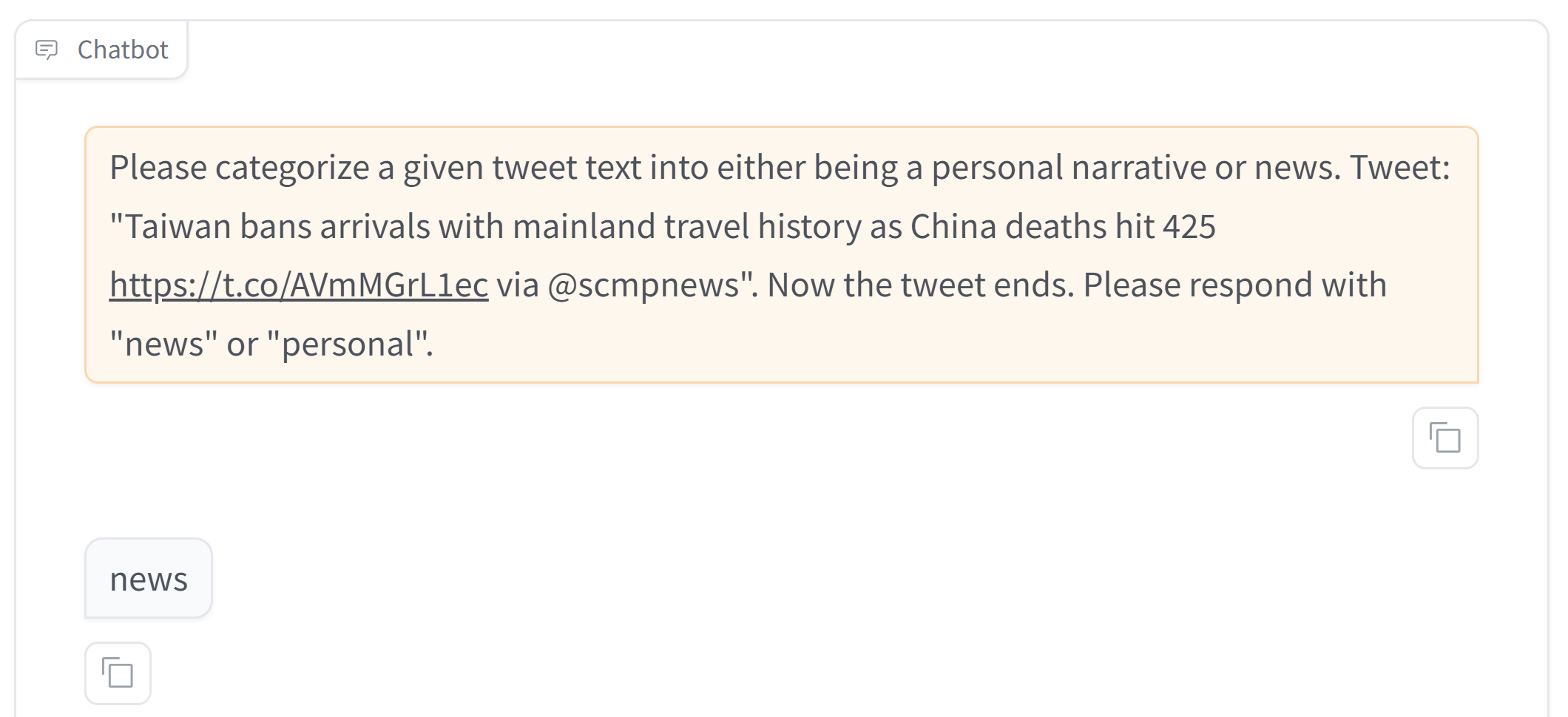

**Public health keywords for filtering the datasets**

public health, global health, health promotion, disease prevention, health education, health policy, health equity, health disparities, access to healthcare, health systems, community health, infectious diseases, chronic diseases, mental health, pandemic, epidemic, outbreak, obesity, diabetes, heart disease, cancer, hiv, aids, tuberculosis, malaria, smoking cessation, alcohol use, substance abuse, healthy eating, physical activity, exercise, nutrition, stress management, sleep hygiene, depression, anxiety, mental health awareness, suicide prevention, mental health services, reducing stigma, air pollution, water quality, sanitation, environmental hazards, climate change, environmental health, vaccine hesitancy, vaccine confidence, vaccination, immunization, herd immunity, vaccine uptake, immunization programs, vaccine safety, health literacy, health communication, misinformation, health campaigns, health promotion strategies, risk communication, digital health, telehealth, e-health, mhealth, health apps, health informatics, health data privacy, wearable technology, universal health coverage, healthcare reform, health insurance, healthcare access, health economics, health services research, healthcare quality, disaster preparedness, emergency response, public health emergencies, pandemic preparedness, contact tracing, maternal health, child health, infant mortality, reproductive health, family planning, prenatal care, breastfeeding, socioeconomic status, education, housing, food security, employment, social support, social determinants of health, health inequality, minority health, indigenous health, health justice, health advocacy, workplace safety, occupational hazards, occupational health, occupational stress, global burden of disease, health in developing countries, international health, global health initiatives, one health, behavior change, health behaviors, behavioral interventions, health belief model

**Languages officially supported by Qwen 2.5**

Arabic, Bengali, Burmese, Cebuano, Chinese, Czech, Dutch, English, French, German, Hebrew, Hindi, Indonesian, Italian, Japanese, Khmer, Korean, Lao, Malay, Persian, Polish, Portuguese, Russian, Spanish, Tagalog, Thai, Turkish, Urdu, Vietnamese

**Instruction-tuning, LoRA, QLoRA, and LoRAPlus**

Instruction-tuning is a technique of training large language models (LLMs) to follow natural language instruction. With instruction-output pairs covering diverse scenarios, LLMs were trained to follow human instruction, even on tasks that are unseen during model training. It has been widely applied to train LLMs in the healthcare domain^37,38^.

Low-Rank Adaptation (LoRA)^39^ introduces small, trainable low-rank matrices $\Delta W$ to the model's weight matrices $W_{o}$. Instead of updating the full weight matrix $W_{o}$, LoRA decomposes the update as: $\Delta W=BA$,where $A\in R^{r\times k}, B\in R^{d\times r}and r\ll min(d,k)$, and update the original weight matrices $W_{o}$ to

$$W_{o}= W_{o}+ \Delta W$$

where, $r$ is the rank, significantly smaller than $d,k$, reducing the number of trainable parameters. During training, $W_{o}$is frozen and does not receive gradient updates, while A and B contain trainable parameters updated with same learning rate. Note both $W_{o}$ and $\Delta W=BA$, are multiplied with the same input, and their respective output vectors are summed coordinate-wise.

Building on this, Quantized Low-Rank Adaptation (QLoRA)^40^ combines quantization with LoRA. The base model weights $W$ are quantized to $W_{q}$ using 4-bit NormalFloat precision to reduce memory usage. Meanwhile, to make sure the model performance is preserved, for the low-rank updates $\Delta W$, it is stored with higher precision using 16-bit BrainFloat.

Low-Rank Adaptation plus (LoRAPlus)^41^ is based on LoRA with separate learning rate setting for matrix $A$ and $B$. For the standard LoRA proposed by Hu et al., the learning rate for A and B is the same. According to Hayou et al, such a setting provably leads to suboptimal learning when embedding dimension is large. Thus, in LoRAPlus, the learning rate of B is set to be $\lambda\times$ that of A, where $\lambda\gg1$ is fixed.

We employ these methods together, which allow efficient fine-tuning of large language models and significantly reducing resource requirements compared to traditional approaches while maintaining performance.

**Evaluation metrics:** $\text{Precision}$**,** $\mathbf{recall}$**,** $\mathbf{F}_{\mathbf{1}}\mathbf{-score}$**, and** $\mathbf{micro}\mathbf{F}_{\mathbf{1}}\mathbf{-score}$

$$\text{precision}=\frac{\mathrm{TP}}{TP+FP}$$

Where $\mathrm{TP}$ is the number of true positives (correctly predicted positive instances), and$FP$ is the number of false positives (incorrectly predicted positive instances).

$$\mathrm{recall}=\frac{\mathrm{TP}}{TP+FN}$$

Where $\mathrm{FN}$ is the number of false negatives (relevant instances that were not retrieved)

Then the $F_{1}-score$ is defined as

$$F_{1}-score=2\times\frac{\text{Precision}\times\text{Recall}}{\text{Precision}+\text{Recall}}$$

This is calculated by focusing on TP, FP, and FN of a specific category, whereas

$${micro F}_{1}-score=2\times\frac{\text{Micro-Precision}\times\text{Micro-Recall}}{\text{Micro-Precision}+\text{Micro-Recall}}$$

where $\text{Micro-Precision}$ and $\text{Micro-Recall}$ are calculated using TP, FP, and FN aggregated across all classes.
